# A set of circulating microRNAs belonging to the 14q32 chromosomic locus identifies two clinically and phenotypically different subgroups of individuals with recent onset Stage 3 type 1 diabetes

**DOI:** 10.1101/2023.10.08.23296650

**Authors:** Guido Sebastiani, Giuseppina Emanuela Grieco, Marco Bruttini, Stefano Auddino, Alessia Mori, Mattia Toniolli, Daniela Fignani, Giada Licata, Laura Nigi, Caterina Formichi, Alberto Pugliese, Carmella Evans-Molina, Lut Overbergh, Timothy Tree, Mark Peakman, Chantal Mathieu, Francesco Dotta, INNODIA investigators

**Author notes:** Contact info: Prof. Francesco Dotta, Diabetes Unit, Dept. of Medicine, Surgery and Neurosciences, University of Siena, Siena, Italy. Shared First co-authorship (Guido Sebastiani, Giuseppina Emanuela Grieco). INNODIA: “Innovative approaches to understanding and arresting type 1 diabetes”- List of contributors included with submission.

## Abstract

Previous research has indicated that circulating microRNAs are linked to the onset and progression of type 1 diabetes mellitus (T1DM), making them potential biomarkers for the disease. In this study, we employed a multiplatform sequencing approach to analyze circulating microRNAs in an extended cohort of individuals recently diagnosed with T1DM from the European INNODIA consortium. Our findings revealed that a specific set of microRNAs located within the T1DM susceptibility chromosomal locus 14q32 distinguishes two distinct subgroups of T1DM individuals. To validate our results, we conducted additional analyses on a second cohort of T1DM individuals, independently confirming the identification of these two subgroups, which we have named Cluster A and Cluster B. Remarkably, Cluster B T1DM individuals, who exhibited increased expression of 14q32 miRNAs, displayed a different peripheral blood immunomics profile, possessed a lower T1DM risk HLA genotype, and showed better glycaemic control during follow-up visits compared to Cluster A individuals. Taken together, our findings suggest that this specific set of circulating microRNAs located in the 14q32 locus can effectively identify T1DM subgroups with distinct characteristics and different clinical outcomes during follow-up.

**GRAPHICAL ABSTRACT:** 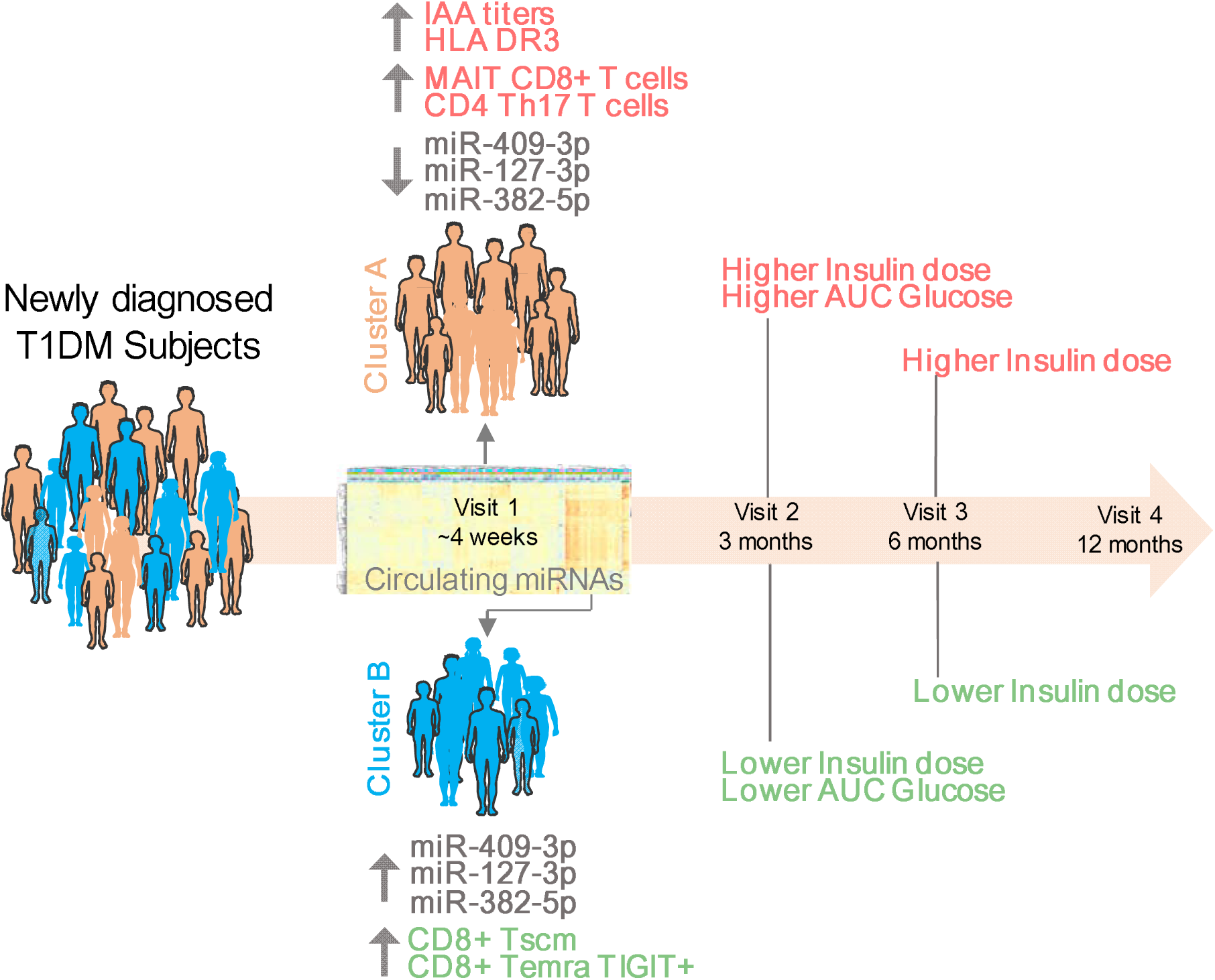

**HIGHLIGHTS:**

- Circulating miRNA profiles in individuals with newly diagnosed Type 1 Diabetes Mellitus (T1DM) can distinguish two subgroups: Cluster A and Cluster B.
- miR-409-3p, miR-127-3p, and miR-382-5p are increased in the plasma of individuals in Cluster B.
- Individuals in Cluster B showed lower IAA titers, a reduced prevalence of HLA risk genotype, and an improved glycaemic profile during the follow-up period.
- Immunomic profiling revealed a reduced frequency of pro-inflammatory immune cells and a higher frequency of exhausted T lymphocytes among individuals in Cluster B.

## INTRODUCTION

Type 1 diabetes mellitus (T1DM) is a chronic autoimmune disease caused by immune-mediated destruction and dysfunction of insulin-producing pancreatic beta-cells, resulting in chronic hyperglycaemia, lifelong insulin therapy, and the occurrence of diabetic vascular complications ^1^. Prodromic stage, disease onset and progression are characterised by marked heterogeneity, leading to an incomplete understanding of T1DM pathogenesis and variable success of interventional therapies ^2^. Age at T1DM diagnosis recapitulates profound differences in genetic predisposition ^3,4^, islet autoantibody appearance ^5,6^, disease clinical onset and presentation ^7^, and beta-cell functional decline progression ^8,9^, with younger individuals showing a severe clinical presentation (i.e. T1DM onset with diabetic ketoacidosis) and a more rapid decline of C-peptide during disease progression. A marked heterogeneity among different individuals is also evident in the characteristics of immune-cell infiltrates in pancreatic islets ^10–12^ and in circulating islet autoantibodies, with a substantial number of individuals lacking any of the presently-known autoantibodies at the time of diagnosis ^13^. In light of this heterogeneity and the high multifactorial nature of T1DM, the existence of multiple distinct subgroups/phenotypes has been hypothesized ^14^. The most significant discoveries are linked to the identification of T1DM Endotype 1 (T1DME1) and T1DM Endotype 2 (T1DME2), which were characterized by differences in pancreatic immune cell infiltration ^15–17^, as well as other phenogroups distinguished by the first time appearance of islet autoantibodies throughout the natural progression of T1D development (i.e. IAA-first, GADA-first)^18,19^. Overall, these studies demonstrated the existence of potentially distinct subgroups of T1DM individuals; however, it is currently unclear how the identification of these subgroups can be beneficial for a specific interventional therapy and how to easily identify them in the clinical practice. Nevertheless, the classification of individuals with T1DM into distinct disease subgroups still remains of high interest and could be beneficial for a precision medicine approach^20^. This classification could be also crucial for the success of interventional immunotherapies^20^. Hence, it is imperative to find easily accessible and measurable biomarkers which are strongly needed to detect and further characterise putative T1DM subgroups^21^. The analysis of circulating biomarkers coupled to unsupervised, data-driven-omics methodologies may help in the unbiased stratification of T1DM individuals, thus to the identification of novel disease endotypes.

MicroRNAs (miRNAs) are a class of small non-coding RNAs, firstly identified in *Caenorhabditis elegans* ^22,23^, and reported to have a critical role in the regulation of gene expression ^24^. They have been associated to the pathogenesis of T1DM ^25^ by mediating the function and dysfunction of beta-cells^26,27^ as well as immune cells^28–31^. Notably, miRNAs also represent an abundant class of blood-based circulating biomarkers^32,33^; as a matter of fact, circulating miRNAs have been documented as mediators of intercellular communication over long distances in multiple organisms, thus exerting influence on the functional behaviour of recipient cells^34^. Hence, the interception of these intercellular messages holds the potential to yield valuable insights into the status of specific diseases and facilitate the characterization of disease dysfunctions that remain incompletely understood^35^.

Numerous investigations have assessed circulating miRNAs in the plasma or serum of individuals with T1DM. Currently, certain miRNAs have consistently shown repeated associations with disease onset (i.e. miR-24-3p^36–43^, miR-146a-5p^37–39,44,45^, miR-375-3p^37,40,46–49^), while some have also been linked to disease progression (i.e. miR-375-3p^37,50^, miR-24-3p^38,50^), particularly in relation to the decline in beta-cell function. While there have been certain specific and promising repeated associations observed between circulating miRNAs and T1DM, it is important to note that many of other findings have not been consistently confirmed in multiple studies. This underscores the variability in miRNA measurements, which may be attributed to the heightened heterogeneity among cohorts of T1DM individuals, pre-analytical variables that can impact sample collection, and the performance of miRNA analytical platforms. Additionally, to date, an unsupervised and unbiased analysis of circulating miRNAs in T1DM individuals, with the aim of stratifying them into multiple subgroups based solely on their circulating expression levels, has not yet been attempted. In light of the limited overlap among different reports and the lack of a miRNA-based unsupervised classification of T1DM individuals, we employed two different high-throughput sequencing platforms to comprehensively and unbiasedly investigate the circulating expression profile of miRNAs^51^ within two large cohorts of recently diagnosed T1DM individuals (within 6 weeks from diagnosis) who were recruited and followed up as part of the INNODIA consortium project^52^. This approach has allowed us to detect and validate two distinct subgroups of T1DM individuals characterised by different expression levels of a set of miRNAs belonging to the 14q32 chromosomal locus and reporting differences in glycaemic control, HLA, and peripheral blood immune cells profiles.

## RESULTS

### T1DM individuals, data collection and miRNAs study design

Circulating microRNAs profiling from blood-derived plasma was performed in two independent cohorts (initial and validation cohort) of individuals enrolled within 6 weeks from clinical diagnosis of stage 3 T1DM in the European consortium INNODIA Natural history study (https://www.innodia.eu)^52^. The schematic of the study design is reported in **Figure 1**.

**Figure 1.**
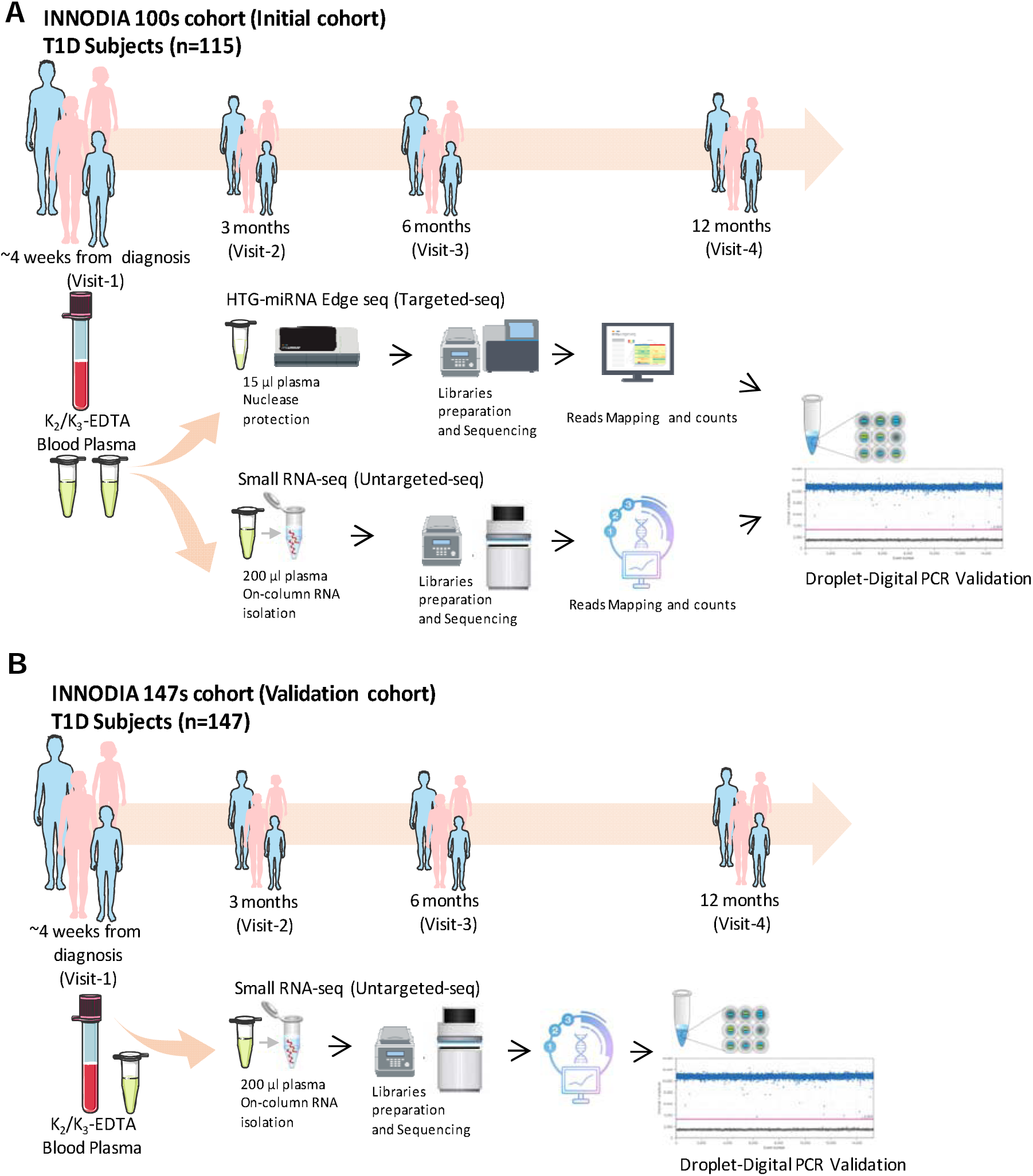
Schematic of the miRNA study design. The study design involved the analysis of two cohorts of Type 1 Diabetes (T1DM) individuals: an initial screening cohort, the INNODIA 100s cohort (**A**), consisting of n=115 T1DM individuals, and a validation cohort, the INNODIA 147s cohort (**B**), consisting of n=147 T1DM individuals. All individuals were followed-up with programmed visits at 3-(visit 2), 6 (visit 3) and 12 months (visit 4) after clinical diagnosis of T1DM. In both cohorts, blood samples were collected at baseline (visit 1) and processed within 2 hours of blood draw. The collected blood samples underwent two consecutive centrifugation steps to separate plasma from contaminant cells and platelets. The plasma samples were then aliquoted (200 μL) and stored at −80°C in a centralized biobank. For the INNODIA 100s cohort, the plasma samples were subjected to miRNA profiling using two different sequencing platforms: HTG-miRNA Edge Seq and Small RNA-seq. The profiling was followed by validation using droplet digital PCR. For the validation cohort, the plasma samples were analyzed using small RNA-seq, and subsequently validated using droplet digital PCR.

The study population of the initial cohort considered for the small RNAs study (here referred to as “100s cohort”) consisted of n=115 recently diagnosed T1DM individuals (sex: 58F/57M; age: 12,4±7,7 years) (mean duration: 4,5 ± 1,5 weeks) (**Table 1**). T1DM individuals were followed-up to 12 months post-diagnosis and subjected to 4 visits (V1: baseline; V2: 3 months; V3: 6 months; V4: 12 months) (**Table S1A**) where the main demographic data, HLA genotype, diabetes related clinical characteristics, and clinical site of recruitment, were measured and collected (**Table 1, Table S1A and S2**). Plasma-EDTA samples for miRNAs analysis were collected at baseline visit (visit 1 or V1) using a standardised protocol adopted by all clinical sites involved in the study^51^. Whole blood samples from a subset of T1DM individuals from the 100s cohort (67/115) were collected at V1, processed to isolate Peripheral Blood Mononuclear Cells (PBMCs), and analysed for circulating immunomic profile, in order to define specific immune cell subsets potentially associated to miRNA expression patterns or T1DM subgroups. Circulating miRNAs from n=115 T1DM individuals were analysed using two different library preparation strategies on all plasma samples, followed by short reads sequencing with Illumina platforms. Specifically, we used: *i*) a RNA extraction-free targeted strategy adopting the HTG Edge-Seq miRNA Whole Transcriptome Sequencing (referred to as “targeted-seq”), and *ii*) a previously standardised untargeted approach^51^ with QIAseq-miRNA/Small RNA Sequencing (referred to as “untargeted-seq”). Hence, each plasma sample was analysed using both methods. Additionally, a subset of plasma samples from n=6 T1DM individuals were run in duplicate for each platform, yielding a total of n=121 samples analysed.

**Table 1.** Baseline clinical characteristics of 100s and 147s cohort of T1DM individuals. Mean values ± standard deviation are reported for continuous variables; n or n% for categorical values. Number of T1DM individuals with available measurements for each specific variable is reported in brackets. BMI-SDS is exclusively reported for T1DM individuals below 18 years of age (100s cohort, n=99)(147s cohort, n=126) at the moment of diagnosis.

| Demographics | 100s cohort | 147s cohort |
| --- | --- | --- |
|  | Baseline (Visit 1) | Baseline (Visit 1) |
| Age (years) | 12.51 ± 7.70 [115] | 12,03 ± 7,82 [147] |
| Sex (Female/Male) | 58/57 | 55/92 |
| BMI (Kg/m<sup>2</sup>) | 23.12 ± 2.82 [16] | 22,26 ± 2,99 [21] |
| BMI_SDS | 0.15 ± 1.09 [99] | 0,38 ± 1,12 [126] |
| Disease duration (weeks) | 4.08 ± 1.53 [115] | 3,91 ± 1,76 [142] |
| Fasting C-peptide (pmol/L) | 277.64 ± 203.54 [115] | 270,03 ± 194,79 [145] |
| HbA1c (mmol/mol) | 77.45 ± 19.44 [112] | 76,01 ± 19,26 [143] |
| Insulin dose (units/kg/day) | 0.52 ± 0.27 [113] | 0,59 ± 0,40 [144] |
| IAA (% positive) | 76 [88] | 77,55 [114] |
| IA2A (% positive) | 72,1 [83] | 78,23 [115] |
| GADA (% positive) | 76 [88] | 77,55 [114] |
| ZnT8A (% positive) | 66 [76] | 70,75[104] |

The study population of the validation cohort (referred to as “147s cohort”) consisted of n=147 T1DM individuals (sex: 55M/92F; age: 11,9 ± 7,9 years) (mean duration: 3,9 ± 1,7 weeks) (**Table 1** and **Table S3A**). Plasma samples of T1DM individuals from 147s cohort were analysed using the untargeted-seq approach and results were further validated through ddPCR for selected miRNAs of interest.

### Circulating miRNAs profile analysis of 100s cohort T1DM individuals using a dual-sequencing approach

Small RNA-seq technologies have enhanced the detection of miRNAs from plasma samples^53^. However, the limited RNA content in plasma, variations in RNA extraction methods, differences in cDNA library preparation protocols and in sequencing approaches have introduced biases into the analytical workflow, resulting in inconsistent findings across various studies^54,55^.

Therefore, to ensure the identification of miRNAs that were consistently detected by both targeted- and untargeted-seq methodologies, we performed a cross-validation of the results obtained from both platforms. This approach allowed us to establish a set of circulating miRNAs that exhibited concordant expression patterns and could therefore be effectively employed for stratifying T1DM individuals at baseline or potentially predicting disease progression during the follow-up period.

In the targeted-seq, two plasma samples failed to generate libraries, resulting in a total of n=119/121 samples sequenced. Overall, sequencing quality metrics including Q30, total yield, and total reads passing filter met the acceptance criteria (**Figure S1A-B**). The mean total read counts for each sample was 4.7 ± 0.88 × 10^6^ reads (**Figure S1C-D)**, while the mean read counts aligned to miRNAs for each sample was 3.2 ± 0.67 × 10^6^ (**Figure S1E**).

In the untargeted-seq, all plasma samples successfully generated cDNA libraries (n=121/121), as shown by the correct sized cDNA fragments analysed by capillary electrophoresis (**Figure S2A-B)**. The quality controls of the untargeted-sequencing metrics returned high quality parameters that met the acceptance criteria, including the *Phred* score (**Figure S2C).** The mean total read counts for each sample was 7.5 ± 2.1 × 10^6^ reads (**Figure S2D-E**), while the miRNAs mean read counts was 2.6 ± 1.3 × 10^6^ (**Figure S2F**). Overall, a total of n=114 unique T1DM individuals were successfully profiled for circulating miRNAs using both sequencing approaches.

Then, we applied a cross-validation strategy to obtain a reliable set of data (**Figure 2A**). Overall, we obtained raw data counts of 2083 miRNAs for the targeted-seq and 2422 miRNAs for the untargeted-seq. After low counts filtering, a total of 892 and 753 miRNAs were retained in the targeted- and untargeted-seq dataset, respectively (**Figure 2A)**. A total of 402 unique miRNAs were commonly detected in both sequencing methods. Next, we selected only those miRNAs that had a positive and significant Pearson’s correlation coefficient for each corresponding miRNA pairs between targeted- and untargeted-seq (**Figure 2B and Figure S3**). By employing this strategy, we successfully obtained two datasets comprising a total of n=248 plasma-derived miRNAs each, that exhibited consistent and concordant expression patterns following cross-validation. Utilizing the expression datasets obtained through the cross-validation approach, we checked the internal platform reproducibility by inspecting the distance matrices of the technical replicates. The analysis of the replicates demonstrated a good internal reproducibility, as the sample duplicates clustered together in both sequencing platforms. (**Figure 2C and Figure S4)**

**Figure 2.**
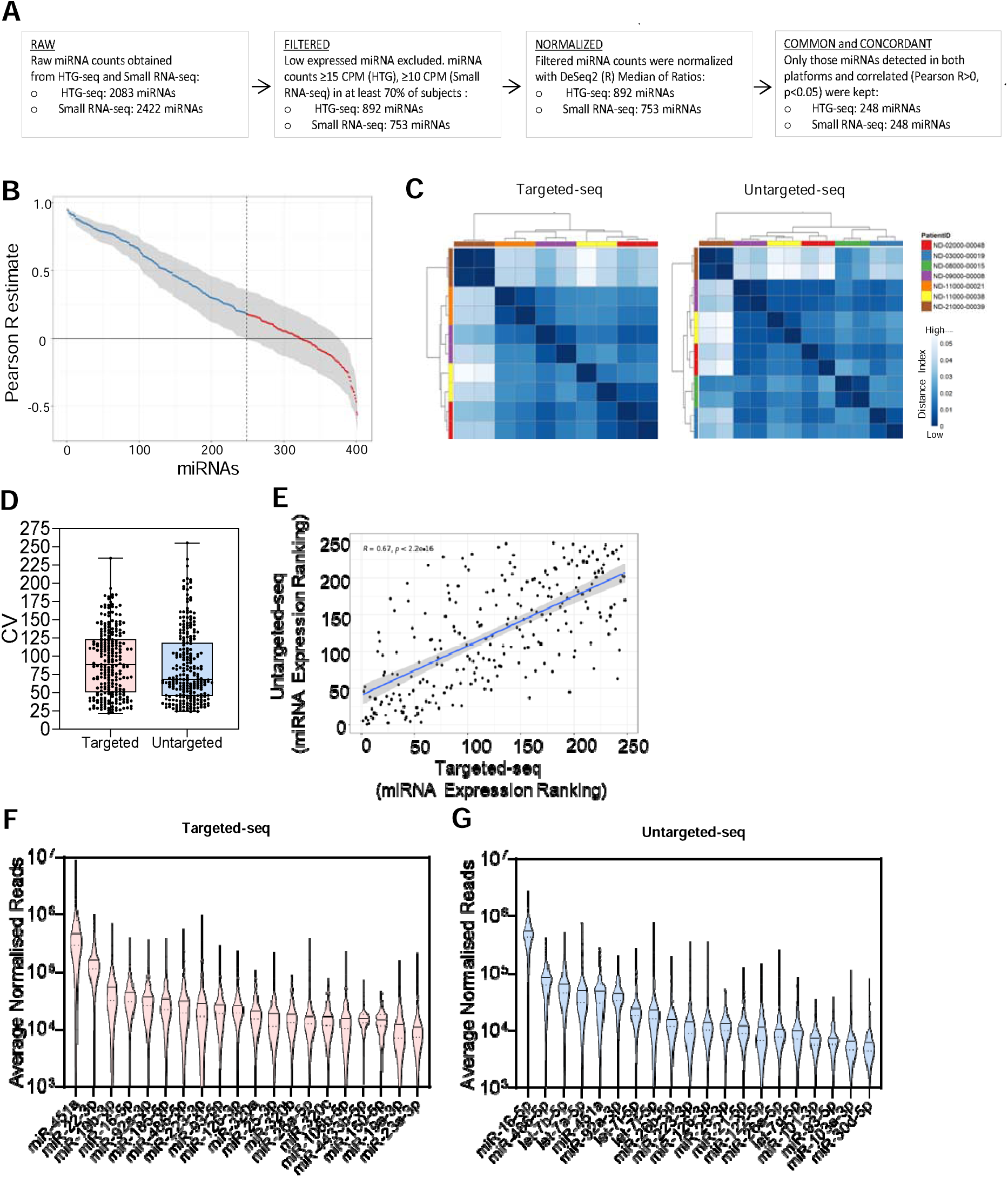
Cross-validation strategy and performance analysis of the dual-platform sequencing approach for plasma samples of T1DM individuals from the 100s cohort. (**A**) Summary of the analytical pipeline for the datasets obtained from the sequencing cross-validation approach using two different platforms on the same set of plasma samples, depicted as a box diagram. (**B**) Correlation analysis using Pearson’s R values on variance-stabilised read counts (rlog transformation from DESeq2 package) reported in the *y* axis, for each commonly detected miRNA (n=402) across the two platforms. Each dot represents a miRNA, and its position on the *x* axis indicates the descending rank of its correlation value between the two platforms, with grey background indicating its 95% confidence interval. Blue dots represent miRNAs with a positive correlation (R-value > 0) and a P-value ≤ 0.05 (n=248), while red dots represent miRNAs with a zero or a negative correlation (n=154, lower confidence limit ≤ 0). (**C**) Sample distance matrix for targeted-seq and untargeted-seq, showing the distance values of technical replicate pairs (n=5 for targeted-seq; n=6 for untargeted-seq) calculated on the n=248 common and concordant miRNAs. The colour scale represents the sample distance calculated as 1-Pearson’s R on rlog transformed reads count, ranging from dark blue (low values indicating similar sample pairs) to white (high values indicating dissimilar sample pairs). (**D**) Coefficient of Variation of miRNA expression (reported as read counts on linear scale) across samples analyzed using targeted-seq and untargeted-seq, focusing on the 248 miRNAs. Each dot represents the coefficient of variation of each given miRNA across all samples analysed in each platform. (**E**) Correlation of miRNA expression ranking between targeted-seq (x-axis) and untargeted-seq (y-axis), evaluated using Spearman’s Rho value for the n=248 miRNAs. (**F**) Top 20 miRNA ranked expression in targeted-seq, reported as the average normalized reads across all analysed samples. (**G**) Top 20 miRNA expression ranking in untargeted-seq, reported as the average normalized reads across all analysed samples. Violin plots in (**F**) and (**G**) report the median values (lines) and quartiles (dotted lines).

The Coefficient of Variation (CV) was calculated for each miRNA read counts across all samples analysed in both platforms. The distribution pattern of CV values showed overlap between targeted sequencing (CV median: 88.1%; 95%CI: 80.8-100.1%) and untargeted sequencing (CV median: 68.7%; 95%CI: 63.7-79.6%). However, there was a tendency towards lower CV values in untargeted sequencing compared to targeted sequencing, as observed in **Figure 2D**. The comparison of miRNA expression levels ranking between targeted- and untargeted-seq demonstrated a significant correlation (**Figure 2E**), indicating a robust performance in miRNA recognition by both platforms. When we specifically examined the 20 most highly expressed miRNAs in the plasma of T1DM patients, we observed a similar top-ranking order between the two platforms, although some exceptions were noted. For example, in the targeted-seq (**Figure 2F**), miR-451a was the most expressed miRNA, while in the untargeted method we detected miR-16-5p as the most expressed one (**Figure 2G**). Overall, the miRNAs expression ranking observed in this study was consistent with previous findings reported in other studies^56,57^.

### Circulating miRNome analysis stratifies T1DM individuals in two subgroups

In order to identify potential subgroups within T1DM individuals using miRNA measurements at baseline, we employed a top-down approach. We conducted unsupervised hierarchical clustering analyses separately for both the targeted and untargeted sequencing datasets **(Figure 3A and 3B**). To determine the number of clusters that optimally define distinct patient subgroups, we applied the silhouette method^58^ on both platforms. This gave us best score for *k* = 2 (**Figure S5**). Thus, by pruning the obtained hierarchical tree with *k* = 2, two distinct groups of T1DM individuals were optimally identified for both targeted-seq (**Figure 3A**) and untargeted-seq (**Figure 3B**). The Principal Component analysis (PCA) confirmed the observed division of T1DM individuals into two subgroups for both platforms (**Figure 3C and Figure 3D**).

**Figure 3.**
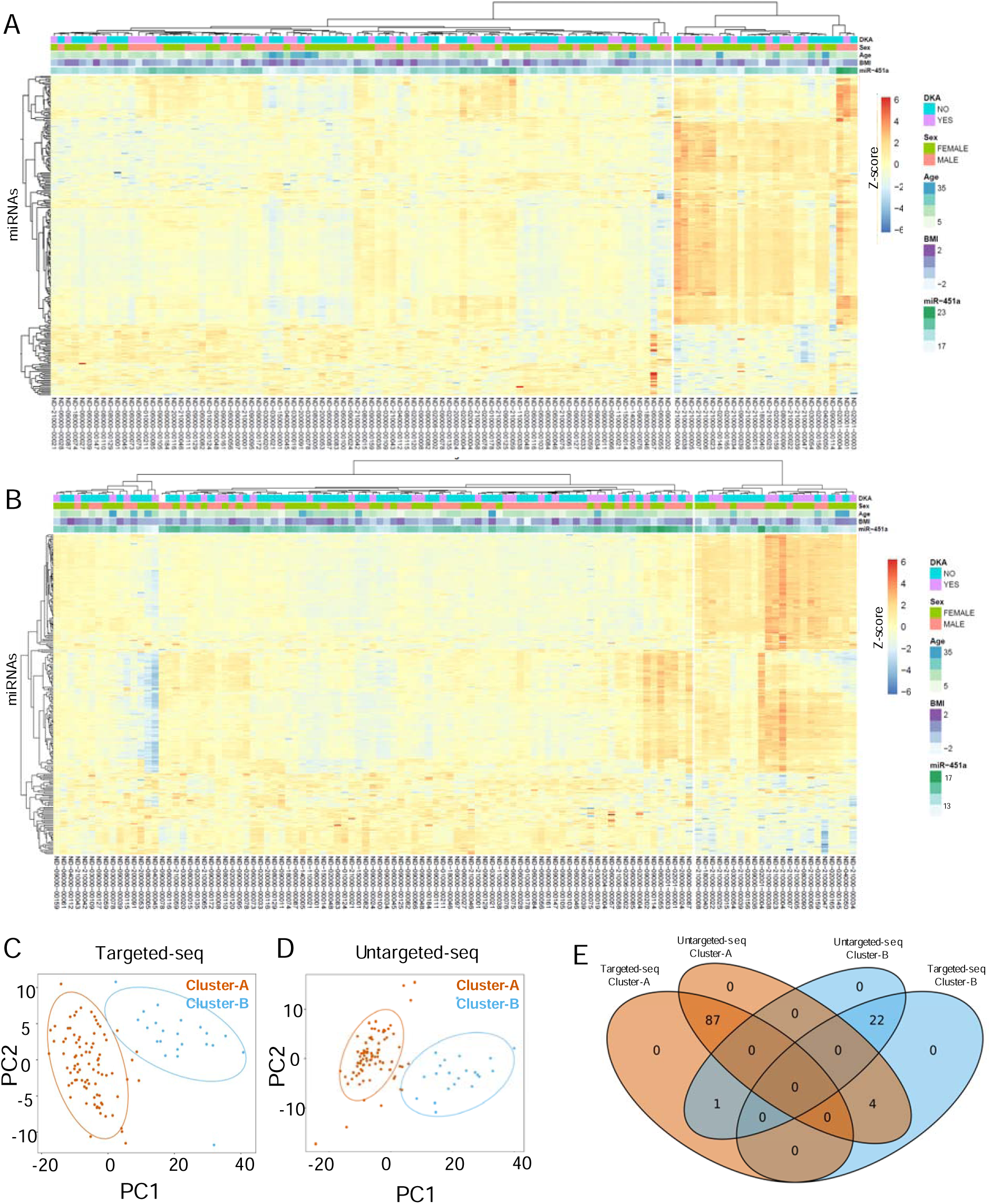
MiRNAs clustering analysis and identification of two distinct T1DM individuals subgroups. (**A**) Unsupervised hierarchical clustering analysis performed on all patients (columns) using Pearson’s R distance on log2 normalized counts (after the addition of a pseudo-count) and complete-linkage as agglomeration method in targeted-seq (**A**) and in untargeted-seq (**B**). The heatmap displays the clustering results, with miRNAs as rows and patients’ information on diabetic ketoacidosis (DKA), sex, age, and the expression of miR-451a (an indicator of haemolysis rate). MiRNA expression is represented as scaled Z-score values ranging from red (+6) to blue (−6). (**C**) Principal Component Analysis (PCA) of the targeted-seq dataset, showing the elliptical grouping of samples based on miRNA expression. (**D**) Principal Component Analysis (PCA) of the untargeted-seq dataset, showing the elliptical grouping of samples based on log2 miRNA expression. (**E**) Venn diagram showing the number of T1DM individuals analyzed using both platforms and consistently identified as belonging to either Cluster-A or Cluster-B by both analytical sequencing methodologies. There were n=87 T1DM individuals in Cluster A and n=22 T1DM individuals in Cluster-B. Additionally, n=5 T1DM individuals were not consistently identified by both platforms.

The two clusters, denoted as Cluster-A and Cluster-B, comprised n=87 and n=22 T1DM individuals, respectively, resulting in a total of n=109 individuals consistently assigned to their respective clusters across both sequencing platforms (**Figure 3E**). The analysis of the clinical site of sample collection (**Figure S6A-B**), the total number of miRNA reads detected in each analytical platform (**Figure S6C-D**), and the haemolysis rate assessed through the erythrocyte-enriched miR-451a (**Figure S6E-F**) did not exhibit significant differences between the two clusters. Hence, these findings suggest that these pre-analytical/analytical variables are not major contributors to the observed clustering.

### Cluster A and Cluster B T1DM individuals showed differences in IAA titres and in T1DM high-risk genotype at baseline

To examine the key clinical parameters potentially associated with Cluster-A and Cluster-B at baseline (visit 1), we firstly fit a logistic regression model including age, sex, BMI (or BMI-SDS), number of autoantibodies/titres, and key metabolic outcomes (**Figure 4A**). In this model, IAA titres at baseline were significantly associated with T1DM subgroups. Of note, the odds ratio of having a higher IAA titre was significantly lower in Cluster B T1DM individuals compared to Cluster A (p=0.0085), Log2 OR: −0.04. 95% CI: −0.009,-0.08; univariate logistic regression analysis) (**Figure 4A and 4B**). Age, sex, BMI (or BMI-SDS), number of autoantibodies, GADA, IA-2A, ZnT8A titres, and other key metabolic outcomes were not significantly associated to these subgroups at baseline (**Figure 4A and Table S1B**). The frequency of T1DM individuals who showed a diabetes onset with ketoacidosis (DKA) (35,5% of total T1DM individuals in 100s cohort) also did not significantly differ between Cluster A (36% with DKA at diagnosis) and Cluster B (46% with DKA at diagnosis) individuals (**Figure S7A and Table S1B**).

**Figure 4.**
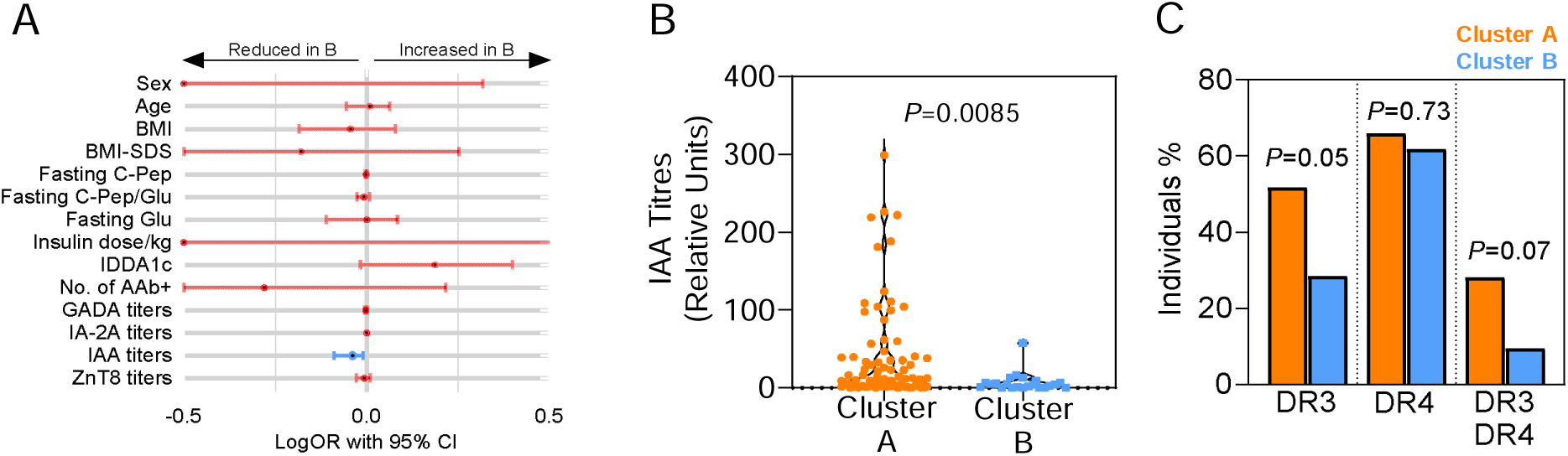
Clinical differences between Cluster A and Cluster B T1DM individuals at baseline (visit 1). (**A**) Forest plot presenting the effect estimates (represented by single dots) and 95% confidence intervals (indicated by bars) for Cluster B across selected clinical variables collected at visit 1. The measure of the strength and direction of the relationship between selected clinical variables and the likelihood of belonging to the Cluster B are presented as log odds ratio. The log odds raio is obtained using univariate logistic regression analysis. Blue bars indicate statistically significant effects (P ≤0.05). (**B**) Comparison of IAA titres at baseline visit 1 in T1DM individuals belonging to Cluster A (n=87) versus Cluster B (n=22); statistical analysis performed using the Mann-Whitney U test (P ≤0.05). (**C**) T1DM risk HLA haplotype distribution in Cluster A and Cluster B individuals; the frequency of the risk genotype was calculated based on available genotype data (107/109 individuals). Chi-square tests were conducted to evaluate the statistical significance of frequency differences between Cluster A and Cluster B (*P* ≤0.05).

HLA risk genotype data, available for 106/109 T1DM individuals, showed that the high-risk HLA genotype DR3-DQ2 [DRB1*03:01-DQA1*05:01-DQB1 *02:01] was more common in individuals from Cluster-A than in those from Cluster-B (A: 51.8% versus B: 28.6%, p=0.05, χ²-test); a similar trend was observed for the DR3/DR4 genotype, although not significant (A: 28.2% versus B: 9.5%, p=0.07, χ²-test) (**Figure 4C**).

### Peripheral blood immunomic profiles of Cluster-A and Cluster-B T1DM individuals showed association with pro-inflammatory and exhausted immune cell phenotype

To look for further potential differences in circulating immune cell subsets between Cluster-A and Cluster-B individuals at baseline, we performed peripheral blood immunomic profile analysis in a subset of T1DM individuals (67/109) belonging to Cluster A and Cluster B (A=48; B=19) (**Figure 5A and 5B**). High-parameter flow cytometry analysis identifed a total of n=150 different immune cell subpopulations (**Figure 5C**) of which 14 were significantly associated with Cluster A or Cluster B (**Figure 5D and Table 2**). Of note, in Cluster B we observed a major significant reduction in the frequency of immune cell subpopulations commonly associated to a proinflammatory context, such as CD8+ CD161+ CD27+ MAIT T cells (decreased in Cluster B versus A, FC=0.44, p=0.0032 (**Figure 5E**) and CD4+Th17 T cells (decreased in Cluster B versus A FC=0.77, p=0.024)(**Figure 5F**), and of those showing the expression of the early activation marker KLRG1, namely CD8+ CD57-KLRG1+ (decreased in Cluster B versus A FC=0.76, p=0.020) and CD8+KLRG1+ (decreased in Cluster B versus A, FC=0.83, p=0.026). Moreover, in Cluster B we observed a significant increase of CD8+CCR7+CD95+CD45RA+ (CD8 Tscm) (**Figure 5G**) and other immune cell subsets showing the expression of the exhaustion marker TIGIT1 (**Figure 5H**).

**Figure 5:**
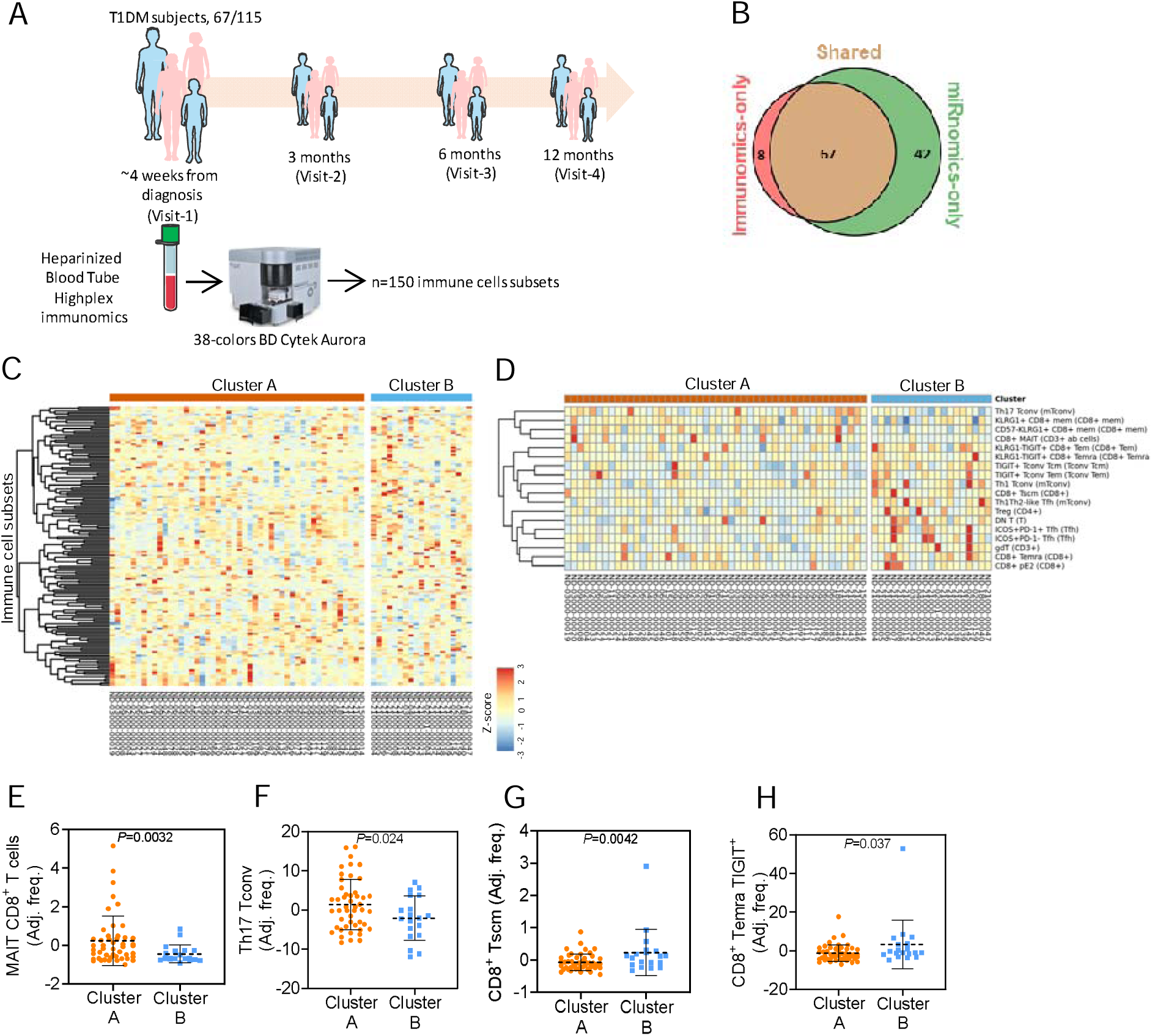
Blood immunomic profile at baseline visit in Cluster A and Cluster B T1DM Individuals. (**A**) Schematic illustration of blood immunomics Analysis of T1DM Individuals at Baseline. The analysis includes the collection of samples and the assessment of n=150 peripheral blood immune cell subsets in a total of n=67 individuals. (**B**) Venn Diagram showing the number of T1DM individuals in the INNODIA 100s Cohort with shared analysis of both miRNomics and immunomics. Among the T1DM individuals, n=67 individuals had complete data available for both miRNomics and immunomics analyses. (**C**) Heatmap and clustering analysis of immune cell subset frequency in Cluster A and Cluster B T1DM individuals. The immune cell subsets demonstrating significant differential frequencies between Cluster A and Cluster B are further illustrated in (**D**) and listed in Table 2. For representative purpose, the proportions of immune cells were residualised with a linear model for the processing effect (same day/overnight) to remove the effect of the covariate and then scaled to Z-score. (**E-H**) Beta regression models of the proportions for selected and relevant immune cell populations. The beta regression depicts the proportions of MAIT CD8+ T cells, Th17 Tconv cells, CD8+ Tscm cells, and CD8+ Temra TIGIT+ cells populations in Cluster A and Cluster B. *P* values (*P* ≤ 0.05) were calculated using a beta regression model correcting for the processing effect; mean and standard deviation (SD) of the proportions of immune cells after residualisation for the processing effect are shown.

**Table 2.** Immune cell subset proportions differentially represented in Cluster B vs Cluster A. The proportions of immune cell subsets were compared between Cluster B and Cluster A, and the results are presented in this table. To facilitate the comparison, the proportions were transformed using the method described by Smithson & Verkuilen^93^, which rescales the dependent variable to the interval (0, 1). The data were analysed using a beta regression model, with correction for the blood processing protocol (same day/overnight). Immune cell populations with a P-value ≤ 0.05, indicating statistical significance, were identified as significantly different between the two groups of patients. The immune cell subsets are listed in order from the most increased to the most decreased in Cluster B compared to Cluster A. The fold change (FC) values indicate the magnitude of the difference. Please note that the direction of change (increased or decreased) refers to Cluster B in relation to Cluster A.

| Immune cell subset | Cluster A | Cluster B | P value | FC (Cluster B vs Cluster A) |
| --- | --- | --- | --- | --- |
| CD8+ TSCM | 0,0039 | 0,0074 | 0,0042 | 1,90 |
| Th1-Th2-like Tfh | 0,0010 | 0,0017 | 0,00008 | 1,69 |
| KLRG1- TIGIT+ CD8+ Temra | 0,0613 | 0,0981 | 0,0374 | 1,60 |
| ICOS+ PD1- Tfh | 0,0215 | 0,0326 | 0,026 | 1,51 |
| ICOS+ PD1+ Tfh | 0,0764 | 0,1143 | 0,0137 | 1,50 |
| CD8 pE2 | 0,0708 | 0,1042 | 0,0217 | 1,47 |
| DN | 0,0469 | 0,0615 | 0,0407 | 1,31 |
| CD8 TEMRA | 0,1916 | 0,2505 | 0,02 | 1,31 |
| KLRG1- TIGIT+ CD8+ Tem | 0,0948 | 0,1169 | 0,0256 | 1,23 |
| Th1 Tconv | 0,1977 | 0,2378 | 0,0248 | 1,20 |
| gdT cells (CD3<sup>+</sup>) | 0,0442 | 0,0528 | 0,0088 | 1,19 |
| TIGIT+ Tconv Tem | 0,1052 | 0,1214 | 0,026 | 1,15 |
| TIGIT+ Tconv Tcm | 0,2682 | 0,2936 | 0,0069 | 1,09 |
| Treg CD4+ | 0,0659 | 0,0710 | 0,0112 | 1,08 |
| KLRG1+ CD8+ mem | 0,6829 | 0,5637 | 0,0264 | 0,83 |
| Th17 Tconv | 0,1583 | 0,1213 | 0,0241 | 0,77 |
| CD57- KLRG1+ CD8+ mem | 0,4674 | 0,3565 | 0,0208 | 0,76 |
| CD8+ MAIT | 0,0127 | 0,0056 | 0,0412 | 0,44 |

Overall, these data indicate a different signature of circulating immune cell subpopulations between the two clusters of T1DM individuals, suggesting a more proinflammatory phenotype in Cluster A and an exhausted phenotype of several immune cell subpopulations in Cluster B.

### Cluster B T1DM individuals have a reduced insulin requirement at follow-up

Next, we investigated whether T1DM individuals in Cluster A and Cluster B had different clinical characteristics during the 12 months of follow-up. Using univariate logistic regression model, we found that individuals in Cluster B were more likely to require a lower insulin dose per kg at 3 and 6 months after diagnosis (**Figure 6A and 6B**), while no differences were observed at 12 months after diagnosis (data not shown). The profile of insulin requirements of T1DM patients in Cluster B, during the follow-up, showed a lower insulin requirement at 3 and 6 months after diagnosis compared to Cluster A, while returning at similar levels at 12 months after diagnosis (**Figure 6C**). These results are independent of the clinical site, as no major differences were found in insulin dosing between the clinical centres involved in the study (**Figure S7B**). Stimulated C-peptide (MMTT AUC) measured at 3, 6 and 12 months showed no significant differences between the two clusters, along with Insulin dose-adjusted HbA1c (IDAA1c) and fasting glucose which did not differ between the two clusters at any visit during follow-up (**Figure 6D, 6E and 6F).** Interestingly, in T1DM individuals from Cluster A, insulin dose and IDAA1c were strongly associated with stimulated C-peptide at V2, as expected, whereas this was not observed in patients from Cluster B (**Figure 6G and 6H**). A similar result was identified at V3, suggesting that the observed difference in insulin dose between the two clusters cannot be fully explained by differences in beta cell function alone. Collectively, these results indicate that additional factors (e.g. peripheral insulin sensitivity) may play a crucial role in this context.

**Figure 6.**
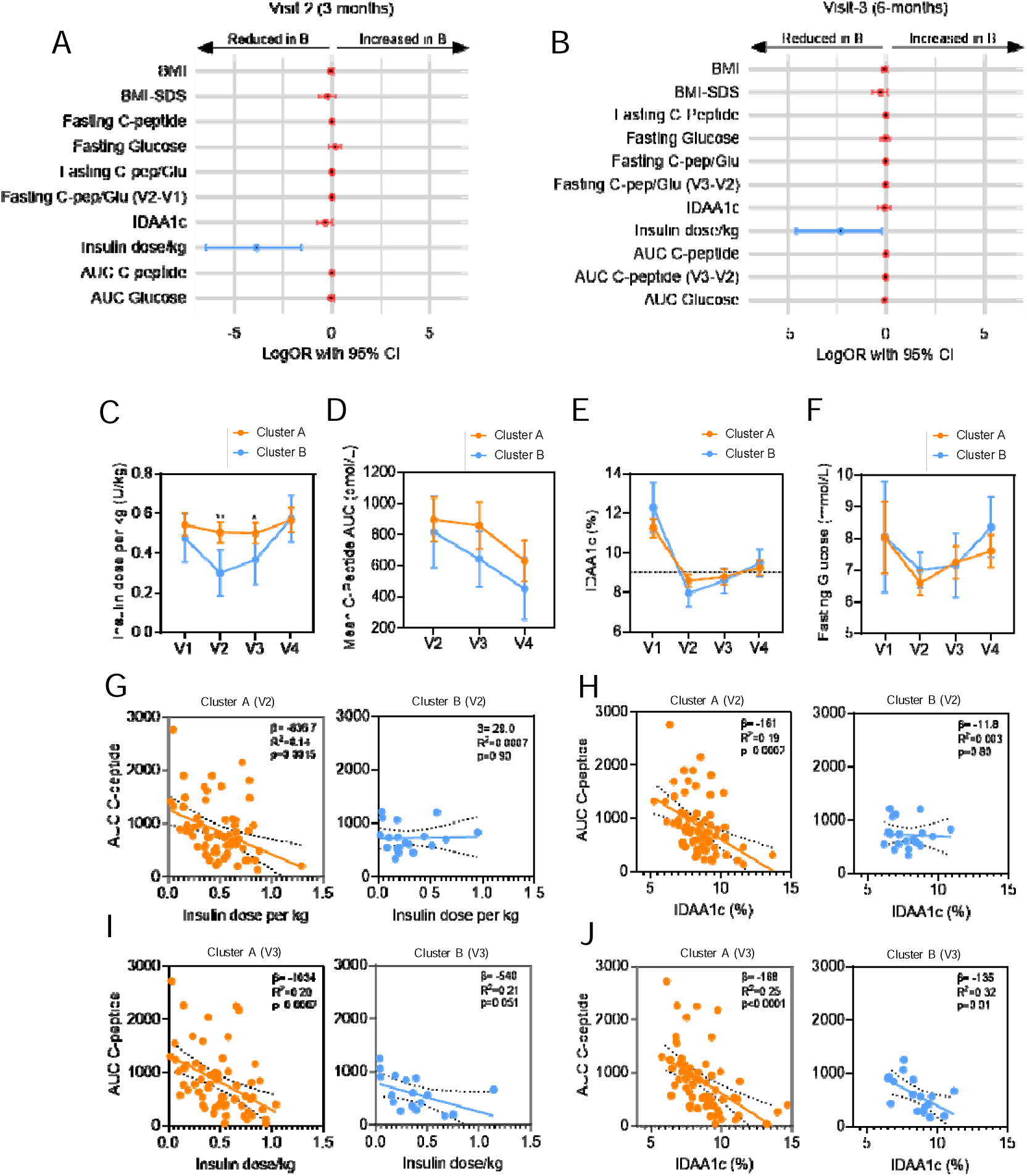
Cluster B T1D individuals showed reduced insulin requirement at follow-up visits. (**A**) Forest plot presenting the effect estimates (represented by single dots) and 95% confidence intervals (indicated by bars) for Cluster B across selected clinical variables collected at (A) visit 2 (3 months) and (**B**) visit 3 (6 months). The measure of the strength and direction of the relationship between selected clinical variables and the likelihood of belonging to the Cluster B are presented as log odds ratio. The log odds raio is obtained using univariate logistic regression analysis.Blue bars indicate statistically significant effects (P ≤0.05). (**C**) Insulin daily dose/kg profile over time from visit 1 (baseline) to visit 4 (12 months) in Cluster A (orange) and Cluster B T1DM individuals; data are reported as median and 95%CI. (**D**) Area under the curve of c-peptide in mixed meal tolerance test (MMTT) performed at visit 2, visit 3 and visit 4 in Cluster A and Cluster B T1D individuals. (**E**) HbA1c-adjusted by insulin dose (IDAA1c) at visit 1, visit 2, visit 3 and visit 4in Cluster A and Cluster B T1DM individuals. (**F**) Fasting glucose at visit 1, visit 2, visit 3 and visit 4 in Cluster A and Cluster B T1DM individuals. Data in (**C**) to (**F**) are reported as median and 95%CI. (**G**) Simple linear regression analysis between AUC C-peptide and insulin daily dose/kg in Cluster A (orange dots) and Cluster B (blue dots) at visit 2 (V2). (**H**) Simple linear regression analysis between AUC C-peptide and IDAA1C in Cluster A (orange dots) and Cluster B (blue dots) at visit 2 (V2). (**I**) Simple linear regression analysis between AUC C-peptide and insulin daily dose/kg in Cluster A (orange dots) and Cluster B (blue dots) at visit 3 (V3). (**J**) Simple linear regression analysis between AUC C-peptide and IDAA1C in Cluster A (orange dots) and Cluster B (blue dots) at visit 3 (V3). Linear regression analyses report slope (β) values, R^2^, and corresponding P values (*P* ≤ 0.05).

### A set of microRNAs belonging to the chromosomic locus 14q32 drives T1DM individuals separation into Cluster A and Cluster B

To comprehensively characterise which set(s) of miRNAs were more relevant for dividing T1DM individuals into Cluster A and Cluster B, we performed a differential miRNA expression analysis between the two clusters. To do this, we analysed the targeted and untargeted seq datasets separately, and then validated the results by selecting only those miRNAs that were significantly differentially expressed with FDR <0.01 on both platforms (**Figure 7A**). In total, we observed n=197 significantly differentially expressed miRNAs between Cluster A and Cluster B. Specifically, n=151 miRNAs were upregulated and n=46 miRNAs were downregulated in Cluster B compared to Cluster A T1DM individuals (FDR <0.01) (**Figure 7A).**

**Figure 7.**
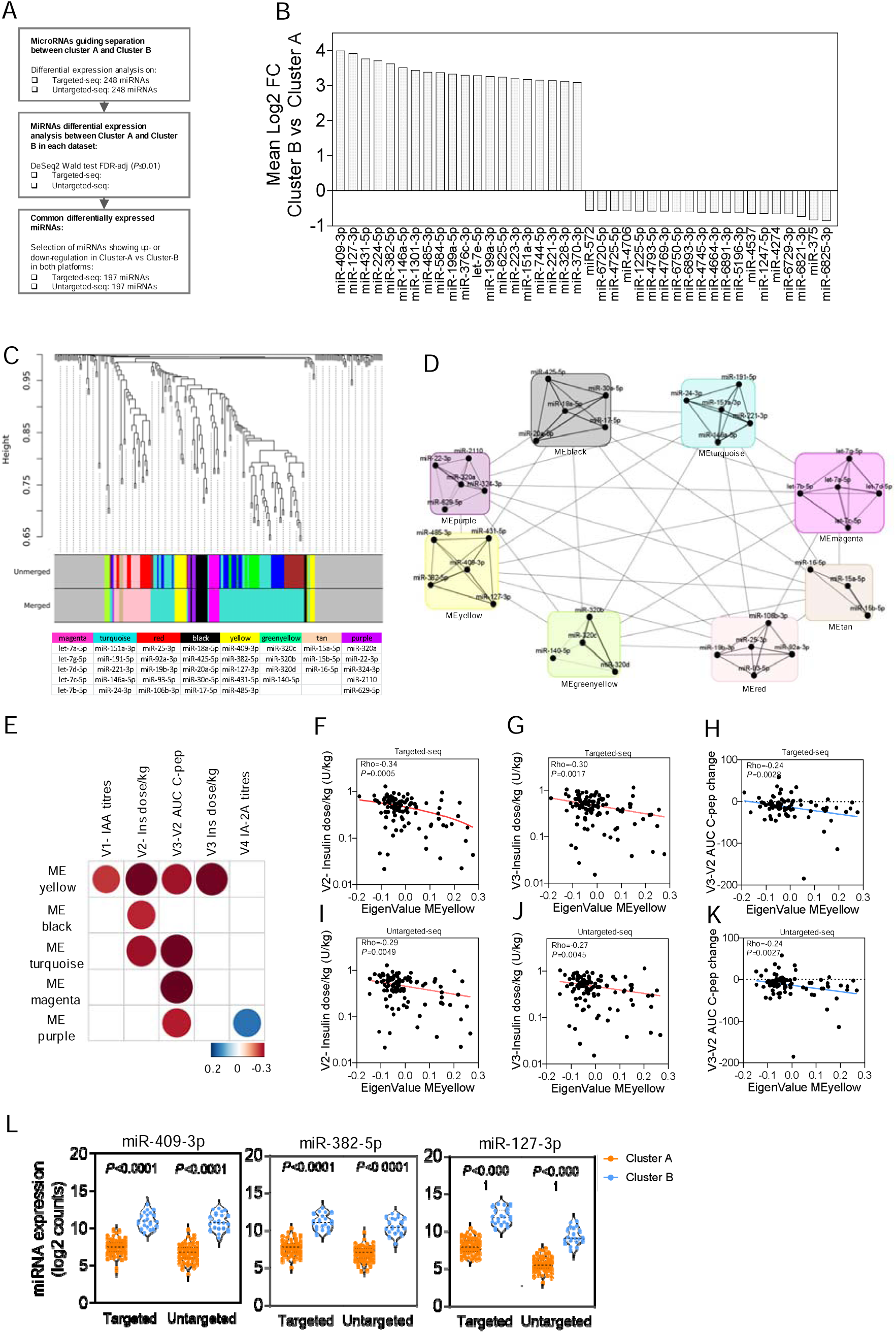
Relevance of microRNAs from the chromosome 14q32 locus in the stratification of T1DM individuals into Cluster A and Cluster B. (**A**) Summary of the analytical pipeline used to determine differentially expressed miRNAs between Cluster A and Cluster B, identified in both targeted and untargeted sequencing datasets. (**B**) Bar plot showing the top 20 differentially expressed miRNAs (up- or down-regulated) in Cluster B vs Cluster A T1DM individuals. Data represents the mean log2 fold change values of Cluster B vs Cluster A individuals obtained from separate analyses of targeted and untargeted datasets. Statistical analysis was performed using the Wald test (DESeq2), considering FDR-adjusted *P* values ≤ 0.01. (**C**) Targeted- and untargeted-seq consensus clustering dendrogram of miRNAs with module colors. The top 5 highly interconnected miRNAs assigned to each module are reported below the hierarchical tree model. (**D**) Scheme illustrating the modules, composition of miRNAs within each module, and the strength of connections among miRNAs. Line thickness in each module indicates the level of interconnectedness among miRNAs. (**E**) Correlation plot based on Spearman Rho values (scale colors from blue to red) obtained between selected clinical parameters at baseline or follow-up visits and eigenvalues of the top 5 miRNAs for each module. Only significant correlations (*P* ≤ 0.05) in both targeted and untargeted-seq are shown, calculated using Spearman’s Rho test. (**F-K**) Scatter plots depicting significant and relevant correlations between eigenvalues of the MEyellow module and clinical parameters for each T1DM subject in the targeted (**F-H**) and untargeted (**I-K**) datasets. Specifically, insulin daily dose/kg at visit 2 (**F, I**), at visit 3 (**G, J**), and changes in area under the curve of C-peptide between visit 2 and visit 3 (**H, K**). Spearman’s Rho test (p ≤ 0.05) was performed, reporting for each graph *rho* and *P* values. (**L**) Comparison of expression levels of miR-409-3p, miR-382-5p, and miR-127-3p in Cluster A vs Cluster B T1DM individuals in targeted- and untargeted-seq. Values are presented as log2 read counts. Statistical analysis using the Wald test (DESeq2) with FDR-adjusted *P* ≤ 0.01.

By ranking the miRNAs from the most upregulated to the most downregulated in Cluster B, we observed a significant enrichment of miRNAs originating from the chromosomal locus 14q32. Specifically, miR-409-3p [chr14: 101065300-101065378 (+), GRCh38] emerged as the most highly upregulated miRNA in Cluster B (Log2 FC: 3.99, B vs A; FDR<0.01), followed closely by miR-127-3p [chr14: 100882979-100883075 (+)] (Log2 FC: 3.92, B vs A FDR<0.01) (**Figure 7B**). Notably, four out of the five most upregulated miRNAs in Cluster B were derived from the 14q32 locus, including miR-409-3p, miR-127-3p, miR-431-5p, and miR-382-5p (**Figure 7B**).

To investigate the presence of modules or groups of closely related miRNAs that may contribute to distinguishing the two clusters or correlate with clinical parameters at baseline or follow-up, we conducted a miRNA network analysis. We employed a weighted miRNA correlation network analysis (WMCNA) to identify distinct miRNA modules. This analysis was performed on both sequencing datasets, and a consensus hierarchical tree model was constructed to identify common miRNA modules across the two analytical platforms. The results of the topological overlap matrix from each dataset, along with the module assignment, are presented in **Figure 7C**. We used a minimum of 3 miRNAs per module and a dissimilarity threshold of 0.1; we identified a total of 8 well-defined eigengene modules of miRNAs, color-coded for easy reference (**Figure 7C**). To identify the most interconnected and relevant miRNAs within each module, we selected the top 5 hub miRNAs and calculated the eigenMiRNA value as a surrogate measure of module expression (**Figure 7D**). Notably, we observed that the yellow module almost exclusively consisted of miRNAs from the 14q32 locus, which were predominantly upregulated in Cluster B compared to Cluster A. Among these miRNAs, miR-409-3p exhibited the highest level of interconnectedness (**Figure 7E**). Subsequently, we examined the correlation between the eigenMiRNA module values and available clinical parameters for each sequencing dataset. The correlation plot analysis, showed in **Figure 7F**, summarizing the significant correlations obtained from both sequencing datasets between modules and clinical parameters at baseline or follow-up. Notably, the yellow module (MeYellow) exhibited a significant association with IAA titers at baseline and with insulin dose per kg at follow-up visits after three (V2) and six months (V3). Additionally, the yellow module displayed a significant inverse correlation with the change in C-peptide levels between visits V2 and V3 (**Figure 7H-K**). The correlation analysis between the yellow module and clinical parameters recapitulates the previously observed differences between Cluster A and Cluster B T1DM individuals. Indeed, those miRNAs showing the highest connectivity in the yellow module (i.e. miR-409-3p, miR-382-5p and miR-127-3p) differed significantly between T1DM patients from Cluster A and Cluster B in both sequencing platforms (i.e. miR-409-3p targeted, Cluster A:7.4 ± 0.95 vs Cluster B: 11,03 ± 1,0; miR-409-3p untargeted, Cluster A: 6,7 ± 1,088 vs Cluster B: 10.9 ± 1,1, p<0.0001; miR-382-5p targeted, Cluster A: 7.6 ± 0.9 vs Cluster B: 11.0 ±0.9; miR-382 untargeted, Cluster A: 6.9 ± 0.8 vs Cluster B: 6.9 ± 0.8, p<0.0001; miR-127 targeted: Cluster A: 7.6 ± 0.9 vs Cluster B: 11.8 ± 1.0; miR-127 untargeted, Cluster A: 5.5 ± 0.9 vs Cluster B: 9.2 ±1.0, p<0.0001 log2 reads counts) (**Figure 7L**) and were therefore selected for a droplet-digital PCR validation analysis.

### Droplet Digital PCR validation of miR-409-3p, miR-382-5p and miR-127-3p as classifiers of Cluster A and Cluster B T1DM individuals

The present findings underscore the discriminatory potential of miR-409-3p, miR-382-5p, and miR-127-3p in distinguishing T1DM individuals from Cluster A and Cluster B. Moreover, we demonstrated their high interconnectedness within a distinct network module (MEyellow), which almost exclusively comprised 14q32 miRNAs. Hence, using droplet digital PCR (ddPCR) we further analysed miR-382-5p, miR-409-3p, and miR-127-3p in plasma samples of T1DM patients derived from Cluster A and Cluster B of 100s cohort. The results confirmed a significantly higher expression level of miR-382-5p, miR-409-3p, and miR-127-3p in Cluster B individuals compared to Cluster A (**Figure 8A-C**). ROC curve analysis demonstrated the high specificity and sensitivity of these miRNAs in assigning T1DM individuals to the identified clusters. Given the significant correlation between the miRNAs in the MEyellow module and insulin requirements at follow-up, we replicated the analysis using the absolute quantification dataset obtained from droplet digital PCR. Notably, all three miRNAs exhibited a significant inverse correlation with the insulin dose per kg at visits V2 (**Figure 8D**) and V3 (**Figure 8E**).

**Figure 8.**
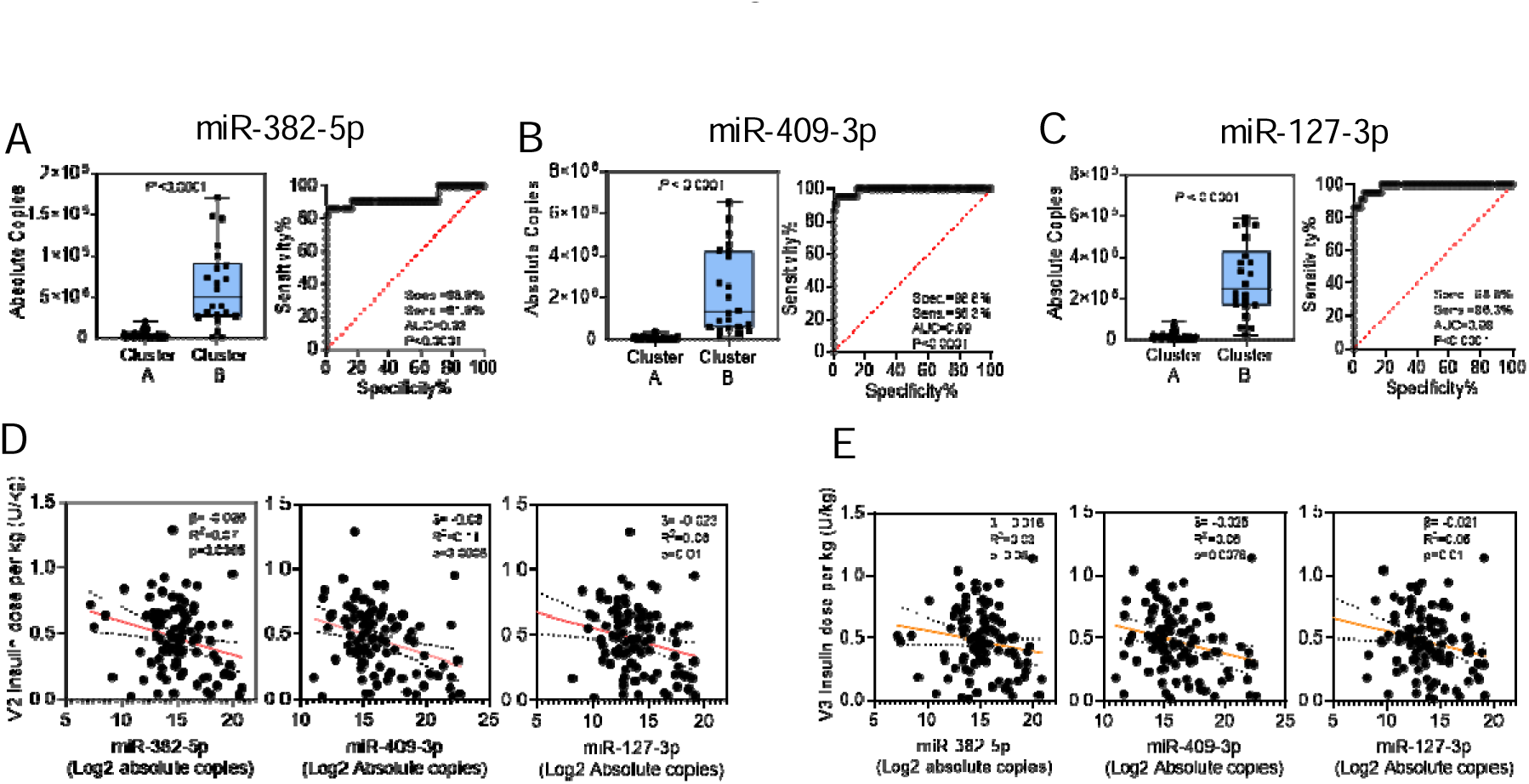
Droplet Digital PCR validation of miR-382-5p, miR-409-3p, and miR-127-3p. (**A**) Stem-loop Reverse Transcriptase and TaqMan-based droplet digital PCR analysis of circulating miR-382-5p in Cluster A and Cluster B T1DM individuals of the 100s cohort. (**B**) Stem-loop Reverse Transcriptase and TaqMan-based droplet digital PCR analysis of circulating miR-409-3p in Cluster A and Cluster B T1DM individuals of the 100s cohort. (**C**) Stem-loop Reverse Transcriptase and TaqMan-based droplet digital PCR analysis of circulating miR-127-3p in Cluster A and Cluster B T1DM individuals of the 100s cohort. Logistic regression and ROC curve analysis for each of the three miRNAs are also presented. Statistical analysis was performed using the non-parametric Mann-Whitney U test (p ≤ 0.05). Logistic regression and ROC curves provide information on specificity, sensitivity, area under the curve, and corresponding P values (p ≤ 0.05). (**D, E**) Simple linear regression analyses between miR-382-5p, miR-409-3p, and miR-127-3p, and visit 2 (V2) insulin daily dose/kg (**D**) and visit 3 (V3) insulin daily dose/kg (**E**). Linear regression analyses report slope (β) values, R^2^, and corresponding P values (*P* ≤ 0.05).

### 14q32 miRNAs distinguish Cluster A and Cluster B T1DM individuals in the 147s validation cohort

To further validate the identification of Cluster A and Cluster B subgroups in T1DM individuals, we conducted an analysis on an additional cohort consisting of n=147 T1DM individuals from the INNODIA consortium (**Table 1, Table S3A and Table S4**). This cohort was enrolled and followed up in a similar manner to the initial 100s cohort. As an internal control, we included a set of plasma samples from eight T1DM individuals of the second cohort, which were run in duplicate. Additionally, we included n=2 batch control plasma samples from the initial cohort to test the inter-platform reproducibility.

Validation cohort samples analysis was performed using the small RNA-seq pipeline previously described (untargeted-seq), followed by ddPCR analysis of relevant miRNAs. The sequencing metrics, as shown in **Figure S8**, confirmed the validity of the sequencing run also in this second cohort.

In the 100s cohort, we have successfully identified a robust set of 248 miRNAs using a cross-validation approach involving two distinct platforms. To ensure the reproducibility of our findings, we applied the same rigorous filtering process to the validation cohort. As a result, we were able to detect a total of 226 out of the initial 248 miRNAs in this independent dataset. To assess the reliability of our small RNA-seq method, we conducted correlation analyses on both internal replicates (Pearson R > 0.77, *P* < 2.2×10^-16^) (**Figure S8E**) and inter-cohort replicates (Pearson R > 0.70, *P* < 2.2×10^-16^) (**Figure S8F**); these analyses yielded positive results, further validating the robustness of our approach.

Next, we analysed this dataset by applying a hierarchical clustering analysis pipeline as done on 100s cohort; interestingly, we observed a distribution of T1DM individuals into two clusters (**Figure 9A**), in line with the findings from the initial cohort and resembling the previously identified Cluster A and Cluster B subgroups (Cluster A=105 T1DM individuals; Cluster B=42 T1DM individuals). Additionally, the principal component analysis endorsed the classification of T1DM individuals into distinct groups (**Figure 9B**), supporting the reliability of miRNAs in classifying T1DM individuals into these two clusters. Differential expression analysis revealed an enrichment of 14q32 miRNAs that were upregulated in Cluster B compared to Cluster A (**Figure 9C-9F**). The upregulation of miR-409-3p, miR-382-5p, and miR-127-3p in Cluster B compared to Cluster A was also confirmed using ddPCR (**Figure 9G-I**), providing additional support to the analysis of the sequencing dataset. Next, we investigated the clinical differences between Cluster A and Cluster B T1DM individuals in the 147s cohort. In line with 100s cohort findings, we did not observe differences in age, sex, BMI (or BMI-SDS), number of autoantibodies, GADA, IA-2A, ZnT8A titres, and other key metabolic outcomes (**Table S3B**). Surprisingly, we did not find any significant differences in terms of IAA titers at baseline or insulin dose at follow-up visits, although a trend was evident for both parameters (**Figure S9A, S9B**). However, we observed a significant reduction in MMTT AUC glucose at the follow-up visit V2 in Cluster B T1DM individuals (Cluster A: 13.5±3.3; Cluster B: 11.5±2.9 mmol/l, p=0.0045), indicating a better glycaemic control compared to Cluster A T1DM individuals at follow-up (V2) (**Figure S9B, S9C**). Importantly, this reduction in MMTT AUC glucose in Cluster B T1DM individuals was independent of beta-cell functional parameters (**Figure S9B**) and consistent with the observations made in the initial cohort.

**Figure 9.**
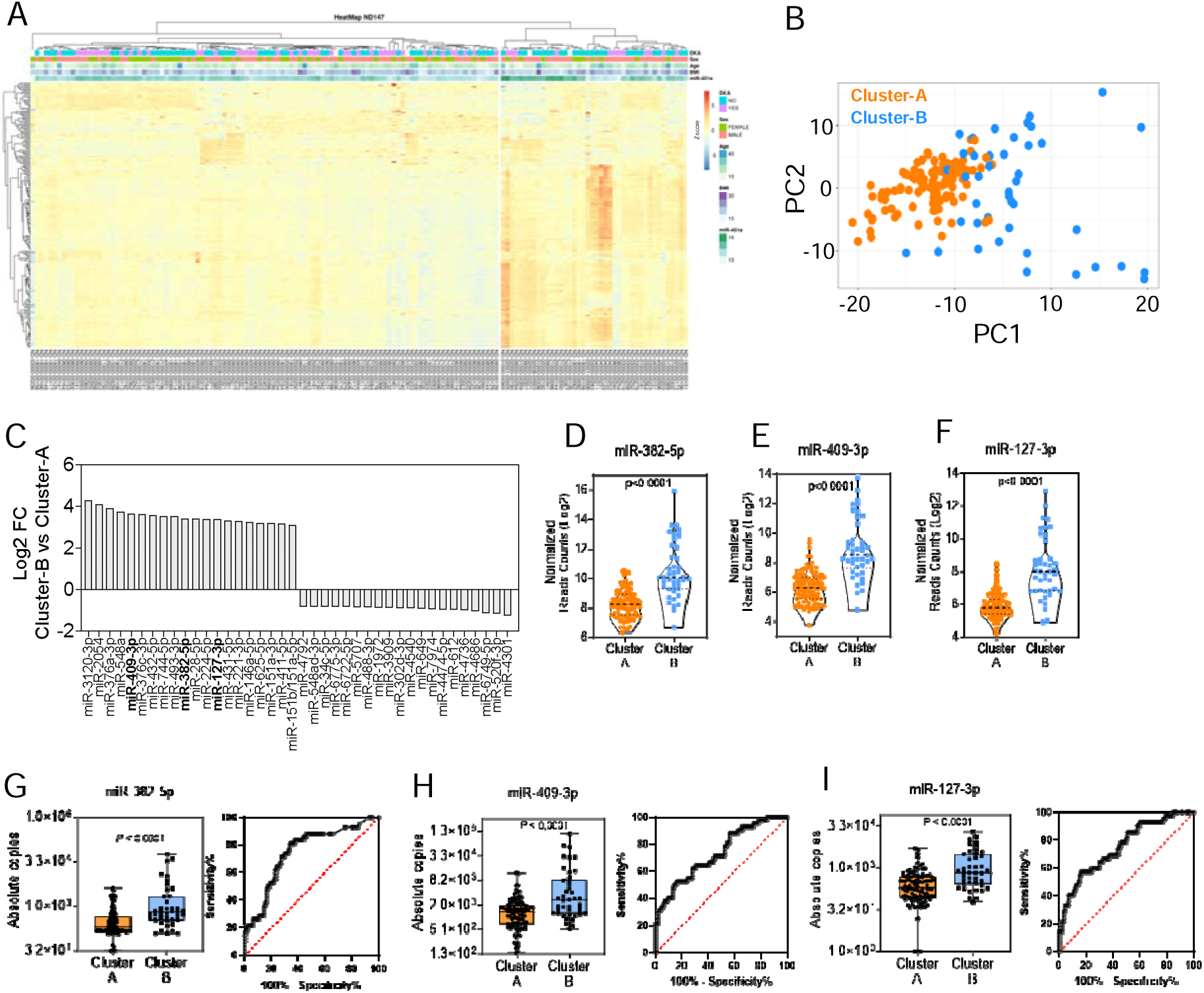
Confirmation of Distinct Clustering of T1D Individuals into Cluster A and Cluster B through circulating microRNA Analysis of the 147s cohort samples. (**A**) Unsupervised hierarchical clustering analysis performed on all patients (columns) using Pearson’s R distance on log2 normalized counts (after the addition of a pseudo-count) and complete-linkage agglomeration method. The heatmap displays the clustering results, with miRNAs as rows and patients’ information on diabetic ketoacidosis (DKA), sex, age, and the expression of miR-451a (an indicator of hemolysis rate). MiRNA expression is represented as scaled Z-score values ranging from red (+6) to blue (−6). (**B**) Principal Component Analysis (PCA) of the untargeted-seq dataset of 147s T1DM individuals cohort, showing the grouping of samples based on miRNA expression (orange dots: Cluster A individuals; blue dots: Cluster B individuals). (**C**) Bar plot showing the top 20 significantly upregulated or downregulated miRNAs in Cluster B vs Cluster A T1DM individuals. Data represents the average log2 fold change values of Cluster B vs Cluster A individuals obtained from the analysis of the untargeted datasets. Statistical analysis was performed using the Wald test (DESeq2), considering FDR-adjusted P values ≤ 0.01. miR-409-3p, miR-382-5p and miR-127-3p are reported in bold. (**D-F**) Comparison of expression levels of miR-382-5p (**D**), miR-409-3p (**E**), and miR-127-3p (**F**) in Cluster A vs Cluster B T1DM individuals in untargeted-seq of 147 cohort samples. Values are presented as log2 values of normalised read counts. (**G-I**) Stem-loop Reverse Transcriptase and TaqMan-based droplet digital PCR analysis of circulating miR-382-5p (**G**), miR-409-3p (**H**) and miR-127-3p (**I**) in Cluster A and Cluster B T1DM individuals of the 147s cohort. Logistic regression and ROC curve analysis for each of the three miRNAs are also presented. Statistical analysis was performed using the non-parametric Mann-Whitney U test (*P* ≤ 0.05). Logistic regression and ROC curves provide information on specificity, sensitivity, area under the curve, and corresponding P values (*P* ≤ 0.05).

In summary, these findings demonstrate that miR-409-3p, miR-382-5p, and miR-127-3p can be utilized to stratify newly diagnosed T1DM individuals into two distinct subgroups characterized by different glycaemic control at follow-up visits after diagnosis.

## DISCUSSION

Numerous studies have consistently demonstrated the association between specific circulating microRNAs (miRNAs) or sets of miRNAs and various aspects of type 1 diabetes mellitus (T1DM), including its onset, progression, and the decline of beta cell function over time^36–44,46–49,59–62^. Within this field, there is a growing appreciation that circulating miRNAs may serve as helpful and reliable biomarkers for T1DM. However, none of the previous studies have taken an unbiased approach to utilize circulating microRNAs for stratifying individuals with T1DM shortly after disease onset, with the aim of identifying novel disease subgroups. Notably, identifying distinct disease subgroups associated with specific phenotypes may shed light on the heterogeneity of T1DM and potentially aid in the stratification of individuals and their assignment to specific interventional immunotherapy^63^.

In the current study we sought to characterise circulating miRNAs in a large cohort of T1DM individuals using a multiplatform sequencing approach coupled to the analysis of two cohorts of T1DM individuals belonging to the European INNODIA study consortium^52^. It is important to highlight that within the consortium, plasma samples were collected and processed uniformly and consistently, following a standardized operating procedure^51^ adopted by multiple clinical centres and specifically designed to minimize pre-analytical biases that could potentially impact the stability and expression of circulating miRNAs^64^. This quality-controlled approach was implemented to ensure the generation of reliable and accurate miRNAs sequencing dataset(s). As a result, the present study possesses a significant advantage in terms of sample collection and the initial screening of miRNAs. Moreover, in the analysis of the initial cohort, two different sequencing approaches were utilized, further strengthening the validity and robustness of the findings. This comprehensive approach enhances the confidence in the observed miRNA profiles and their potential implications.

Using this approach, we have discovered a panel of three miRNAs (miR-409-3p, miR-382-5p, miR-127-3p) that can effectively differentiate between two distinct groups of individuals with T1DM shortly after disease onset, referred to as Cluster A and Cluster B.

At the initial visit (visit 1), which occurred approximately 4 weeks after disease onset where the miRNA profiles were analysed, we observed that Cluster B individuals displayed significantly lower levels of IAA titres and a lower prevalence of the high-risk T1DM genotype HLA-DR3. In children, the appearance of IAA as the first autoantibody was associated with an increased risk of developing future multiple autoantibodies and of T1DM onset^65^. Moreover, IAA and DR3/DR4 HLA risk haplotype have been previously associated to a rapid progression from stage 2 to stage 3 T1D and also to a rapid decline of beta-cell function^66^. These findings suggest a more severe phenotype of T1DM in Cluster A in respect to Cluster B individuals despite the absence of age differences at diagnosis between the two subgroups, which has been reported to be associated to disease severity. These results were partially replicated in the validation cohort (147s cohort), where we observed a tendency (although not significant) for increased IAA titres at baseline among Cluster A individuals. Furthermore, at visit 2 and visit 3 (3-and 6-months after onset), we observed a notable improvement in the glycaemic profile among individuals in Cluster B compared to Cluster A, underscoring a less severe phenotype of Cluster-B subjects during follow-up. This improvement was consistently observed in both the 100s and 147s cohorts. Specifically, individuals in Cluster B exhibited lower insulin dose per kilogram (in 100s cohort) and a reduced area under the curve (AUC) for glucose during the mixed meal tolerance test (MMTT) (in 147s cohort) when compared to those in Cluster A. These findings suggest that Cluster B T1D individuals had better glycaemic control with more favourable glucose metabolism and are concordant with the less severe phenotype observed in Cluster B individuals at baseline. Interestingly, in both T1DM cohorts, we did not detect any differences in beta-cell functional profiles between Cluster A and Cluster B, measured by fasting or MMTT-stimulated C-peptide, during baseline or follow-up visits. These observations indicate the possible presence of variations in insulin sensitivity between the two clusters of individuals. However, it is important to note that, in these T1DM cohorts, direct measurements of insulin sensitivity were not performed at baseline, and the follow-up visits were influenced by the administration of insulin therapy, potentially impairing a reliable insulin sensitivity measures even if using insulin sensitivity surrogate markers. Due to these limitations, additional focused analyses are necessary to gain a better understanding of the role of insulin sensitivity in delineating the distinctions between the two T1D clusters.

To gain further insights into specific differences between Cluster A and Cluster B, we had the valuable opportunity to investigate the peripheral blood immune cell profiles (immunomics) in a subset of T1DM individuals (n=67/115) from the 100s cohort and to establish a connection between their cluster membership and immune cell characteristics at baseline. Of note, Cluster A individuals exhibited a more pro-inflammatory phenotype with a prominent increased frequency of MAIT CD8+ and Th17 Tconv. This implies an increased activation or dysregulation of immune cells in Cluster A, promoting inflammation and potentially contributing to the progression of T1DM. Intriguingly, IL-17 and MAIT T cells, both increased in Cluster A individuals, were previously associated to inflammatory-based insulin resistance mechanisms by disrupting insulin signalling in multiple target tissues^67,68^. On the other hand, Cluster B individuals showed increased frequency of several immune cell subpopulations exhibiting a partial exhausted phenotype (i.e. increased frequency of CD8+ TIGIT+ T cells) and increased frequency of CD8+ T stem cells memory (Tscm) cells. The role of inhibitory receptors is well known, with TIGIT expression representing a hallmark of T cell exhaustion^69,70^; notably, TIGIT also characterises partially exhausted CD8+ T cells accumulating in teplizumab-responder T1DM individuals after treatment^71,72^. In addition, CD8+ T cell exhaustion also characterizes T1DM individuals experiencing a slow progression of the disease after onset^73^.

On the other hand, the function of Tscm cells in autoimmune diseases is still to be fully deciphered. Overall, Tscm play a crucial role in promoting antitumor and immune reconstitution because of their enhanced stem cell-like self-renewal capacity and can serve as a reservoir of effector T-cells. Circulating Tscm showed increased frequency in T1DM individuals, thus potentially promoting autoimmunity^74,75^. Hence, their precise role in Cluster B T1DM individuals should be further analysed. Collectively, these findings suggest that Cluster A and Cluster B are characterised by distinct immune system features, in line with differences observed between the two clusters. Further research is needed to fully comprehend the underlying mechanisms and clinical implications of these immune cell subpopulations in Cluster A and Cluster B individuals.

In both analysed T1DM cohorts (100s, n=109 T1D subject; 147s, n=147 T1D individuals), the levels of circulating miR-409-3p, miR-382-5p, and miR-127-3p were significantly elevated in the plasma of individuals belonging to Cluster B compared to Cluster A. These miRNAs are the top ones that mostly influenced Cluster A and Cluster B separation, thus emphasizing their potential role in distinguishing the two T1D clusters. Notably, these miRNAs are part of the 14q32 chromosomal locus^76^. Intriguingly, miRNA network analysis showed that these three miRNAs were the most interconnected among others within a specific module (ME Yellow). Remarkably, 13 out of 14 miRNAs present in this module were derived from the 14q32 locus. These findings highlight the significance of this locus, which has been previously indicated as a susceptibility region for T1D^77^. Furthermore, network analysis revealed the presence of additional modules containing miRNAs that have been previously associated with T1D, beta-cell function, or diabetic complications in multiple studies. For example, the magenta module comprised miRNAs from the let-7 family, which have been linked to microvascular complications in diabetes^78,79^; the red and turquoise modules contained miRNAs associated with T1D, inflammation, and beta-cell function, such as miR-93-5p^80^, miR-25-3p^36^, and miR-106b-3p^81^ in the red module and miR-151a-3p^82^, miR-24-3p^36–43^, and miR-146a-5p^82–86^ in the turquoise module. Among these, miR-25-3p displayed the highest degree of interconnectedness within the red module, while miR-151a-3p held this distinction in the turquoise one. Interestingly, both miRNAs have previously been linked to metabolic impairment and beta-cell function in T1D^36,82^. Overall, these findings provide additional support to the validity and robustness of our analysis, as the observed interconnected miRNAs in these modules align with established associations in T1D, inflammation and beta-cell dysfunction. Chromosome 14q32 locus hosts one of the largest polycistronic miRNAs cluster in mammals. In humans, it contains 54 miRNA genes included between DLK1 and DIO3 genes region. This locus is subjected to paternal or maternal differential methylation thus expressed from maternal or paternal inherited chromosome rather than showing a biallelic expression. Hence, the entire miRNAs cluster along with MEG3, RTL1as and MEG8 are maternally expressed, while DLK1, RTL1 and DIO3 are paternally expressed^87^. While it is challenging to definitively determine the origin of the differential expression of circulating miRNAs, several studies have demonstrated that miRNAs from the chromosome 14q32 locus, including miR-409 and miR-127, exhibit high expression and enrichment in human islets compared to other tissues^88^. Furthermore, they are specifically expressed in beta-cells rather than alpha-cells^89–91^; indeed, among the top 10 miRNAs that show higher enrichment in beta-cells compared to alpha-cells, 8 of them are derived from chromosome 14q32^91^.

In addition, it has been shown that these miRNAs are differentially expressed in islets of type 2 diabetic donors and regulated by a differential methylation pattern in the MEG3 Differentially Methylated Region (DMR)^90^. Of note, 14q32 miRNAs are critical to beta-cell function and were reported to regulate a set of target genes encoding specific T1D autoantigens (i.e. miR-409 – PTPRN2 and GAD2 genes)^92^.

The putative association between 14q32 miRNAs and their involvement in T1DM pathogenesis is further bolstered by the observation of a specific single nucleotide polymorphism (SNP) that has been previously identified as associated with T1DM in a Genome-wide Association Study (GWAS)^77^. The presence of this SNP (rs941576), which has been linked to T1DM, provides additional evidence supporting the potential role of the 14q32 locus in the development and progression of the disease.

Additional evidence supporting the role of the 14q32 miRNAs in T1DM derives from studies conducted in the Non-Obese Diabetic (NOD) mouse model. In a previous study^30^, we demonstrated that miR-409-3p expression is decreased in the plasma of diabetic NOD mice in comparison with non diabetic normoglycaemic NOD mice, and this reduction is also observed in the pancreas-infiltrating lymphocytes of the same mice. Moreover, a reduced expression of miR-409-3p was also observed in the plasma of recent onset (<2 years from diagnosis) T1D individuals compared to non-diabetic controls^30^. Interestingly, we found a similar reduction in miR-409-3p expression in Cluster A T1D individuals compared to Cluster B. This reduction in miR-409-3p expression is in line with the more severe phenotype observed in Cluster A individuals, both at baseline and during follow-up visits. These findings suggest that miR-409-3p may play a significant role in T1DM pathogenesis and could serve as a potential biomarker for disease severity. The consistent reduction of miR-409-3p expression in both the NOD mouse model and T1DM individuals strengthens the validity of our present observations.

In summary, the results of this study provide evidence supporting the use of miR-409-3p, miR-382-5p, and miR-127-3p as effective markers for stratifying newly diagnosed individuals with T1DM into two distinct subgroups which display different immune-related characteristics at baseline and different levels of glycaemic control over time. Hence, this stratification can be taken into consideration to verify the responsiveness of T1DM individuals of Cluster A and Cluster B to multiple immunotherapies, thus implementing a tailored precision medicine approach to treat T1DM.

### Limitations of the study

We acknowledge limitations of this study. Firstly, the sample size used for the association study between T1DM subject clusters (A or B) and peripheral blood immunomics was relatively small, consisting of n=67 individuals, while a large number of immune cell subpopulations (n=150) were analyzed. Consequently, we did not perform multiple testing corrections to avoid type 2 errors. Therefore, these results should be considered preliminary and exploratory, requiring further validation in additional studies.

Secondly, in 147s cohort we acknowledge a sex dysbalance, being females more prevalent than males; moreover, several analyses in this cohort are still pending due to the ongoing INNODIA study. These include genotyping and immunomics. Nevertheless, the confirmation of the two clusters in this independent cohort supports the findings of the 100s cohort regarding miR-409-3p, miR-127-3p, and miR-382-5p.

Finally, although the circulating miRNAs were validated using droplet digital PCR (ddPCR) in two different T1DM cohorts, comprising a total of 256 T1DM individuals analyzed, we have yet to determine the cellular origin of these miRNAs and then formulate mechanistic insights of these findings. Hence, additional analyses are required to elucidate their origin and the potential molecular mechanisms involved in their dysregulation, which may occur in one or multiple target organs.

## Acknowledgements

The work is supported by the Innovative Medicines Initiative 2 (IMI2) Joint Undertaking under grant agreement No.115797-INNODIA and No.945268 INNODIA HARVEST. This joint undertaking receives support from the Union’s Horizon 2020 research and innovation programme and EFPIA, JDRF and The Leona M. and Harry B. Helmsley Charitable Trust. GS is supported by University of Siena within *F-CUR* funding program Grant No. 2268-2022-SG-PSR2021-FCUR_001. FD was supported by the Italian Ministry of University and Research (2017KAM2R5_003). This work is also supported by EU within Italian Ministry of University and Research (MUR) PNRR “National Center for Gene Therapy and Drugs based on RNA Technology” (Project No. CN00000041 CN3 Spoke #5 “Inflammatory and Infectious Diseases”. This work is also supported by JDRF and The Leona M. and Harry B. Helmsley Charitable Trust for the project “Collaborative Effort to Identify and Validate miRNA as Biomarkers of T1D”.

## Author Contributions

GS and GEG conducted the analyses, prepared tables and figures, interpreted the data and co-wrote the manuscript. MB, SA, AM and MT analysed the data, prepared the tables and figures, and co-wrote the manuscript. DF, GL, LN, CF evaluated and interpreted the data. AP and CEM participated in the design of the study. LO, TT, MP, CM, initiated, designed and supervised the study. GS and FD designed, supervised the study and wrote the manuscript. GS, GEG and FD are the guarantors of this work and, as such, had full access to all the data in the study and takes responsibility for the integrity of the data and the accuracy of the data analysis.

## Data Availability and Code availability

The generated data is person-sensitive and access can be provided by application to the INNODIA Data Access Committee. Any additional information regarding the data reported in this paper is available from the lead contact upon request.

## ^§^Members of the INNODIA and INNODIA HARVEST consortia

Mathieu C, Gillard P, Casteels K, Overbergh L (KU Leuven, Belgium), Dunger D, Wallace C, Evans M, Thankamony A, Hendriks E, Bruggraber S, Qureshi A, Marcovecchio L, Paediatrics laboratory staff (University of Cambridge, UK), Peakman M, Tree T (King’s College London, UK), Morgan N, Richardson S (University of Exeter, UK), Todd J, Wicker L (University of Oxford, UK), Mander A, Dayan C, Alhadj Ali M (Cardiff University, UK), Pieber T (Medical University of Graz, Austria), Eizirik D, Cnop M (Universite Libre de Bruxelles, Belgium), Brunak S (University of Copenhagen, Denmark), Pociot F, Johannesen J, Rossing P, Legido Quigley C (Herlev University Hospital, Region Hovedstaden, Denmark), Mallone R, Scharfmann R, Boitard C (Cochin Institute Paris, France), Knip M, Otonkoski T (University of Helsinki, Finland), Veijola R (University of Oulu, Finland), Lahesmaa R, Oresic M, Toppari J (University of Turku, Finland), Danne T (Children’s and Youth Hospital Hannover, Germany), Ziegler AG, Achenbach P, Rodriguez-Calvo T (Helmholtz Zentrum Muenchen, Germany), Solimena M, Bonifacio E, Speier S (TU Dresden, Germany), Holl R (University of Ulm, Germany), Dotta F (University of Siena, Italy), Chiarelli F (University of Chieti, Italy), Marchetti P (University of Pisa, Italy), Bosi E (University Vita-Salute San Raffaele, Italy), Cianfarani S, Ciampalini P (Bambino Gesù Children’s Hospital, Italy), de Beaufort C (Centre Hospitalier de Luxembourg, Luxemburg), Dahl-Jørgensen K, Skrivarhaug T, Joner G, Krogvold L (Oslo University Hospital, Norway), Jarosz-Chobot P (Medical University of Silesia, Poland), Battelino T (University of Ljubljana, Slovenia), Thorens B (University of Lausanne, Switzerland), Gotthardt M (Radboud University Medical Center, the Netherlands), Roep B, Nikolic T, Zaldumbide A (Leiden University Medical Center, the Netherlands), Lernmark A, Lundgren M (Lund University, Sweden), Costecalde G (Univercell-Biosolutions, France), Strube T, Schulte A, Nitsche A, (Sanofi, Germany), Peakman M, Vela J (Sanofi, United States), von Herrath M, Wesley J, (Novo Nordisk, Denmark), Napolitano-Rosen A (GlaxoSmithKline, UK), Thomas M, Schloot N (Eli Lilly, UK), Goldfine A, Waldron-Lynch F, Kompa J, Vedala A, Hartmann N, Nicolas G (Novartis Pharma AG, Switzerland), van Rampelbergh J, Bovy N (Imcyse SA, Belgium), Dutta S, Soderberg J, Ahmed S, Martin F, Latres E (JDRF, USA), Agiostratidou G, Koralova A (The Leona M. and Harry B. Helmsley Charitable Trust, USA).

## Associated clinical sites

Willemsen R (Barts Health NHS Trust, UK), Smith A (Northampton General Hospital NHS Trust, UK), Anand B (West Suffolk NHS FT, UK), Puthi V (North West Anglia NHS FT, UK), Zac-Varghese S (East & North Hertfordshire NHS Trust, UK), Datta V (Norfolk & Norwich University NHS FT, UK), Dias R (Birmingham Women’s and Children’s NHS FT, UK), Sundaram P (University Hospitals of Leicester NHS Trust, UK), Vaidya B (Royal Devon & Exeter NHS FT, UK), Patterson C (NHS Fife, UK), Owen K (Oxford University Hospitals NHS FT, UK), Dayan C (Cardiff & Vale University Health Board, UK), Piel B (Queen Elizabeth Hospital, King’s Lynn FT, UK), Heller S (Sheffield Teaching Hospitals NHS FT, UK), Randell T, Gazis T (Nottingham University Hospitals NHS Trust, UK), Bismuth Reismen E, Carel J-C (Hospital Robert Debre, France), Riveline J-P, Gautier J-F (Hospital Lariboisiere, France), Andreelli F (Hospital Lapitie-Salpetriere, France), Travert F (Hospital Bichat Claude Bernard, France), Cosson E (Hospital Jean-Verdier, France), Penfornis A, Petit C (Centre Hospitalier Sud-Francilien, France), Feve B (Hospital St Antoine, France), Lucidarme N (Hospital Jean-Verdier Pediatrie, France), Cosson E (Hospital Avicenne, France), Beressi J-P (Hospital Andre Mignot, France), Ajzenman C (Hospital Andre Mignot Pediatrie, France), Radu A (Hospital Europeen Georges-Pompidou, France), Greteau-Hamoumou S (Hospital Louis Mourier, France), Bibal C (Hospital Kremlin Bicetre, France), Meissner T (Universitatsklinikum der Heinrich-Heine-Univeritat Dusseldorf, Germany), Heidtmann B (Katholisches Kinderkrankenhaus Wilhelmstift, Germany), Toni S (AOU Meyer, Italy), Rami-Merhar B (Medical University of Vienna, Austria), Eeckhout B, Peene B, Vantongerloo N (Algemeen Ziekenhuis Geel Sint-Dimpna Geel, Belgium), Maes T, Gommers L (Imeldziekenhuis Bonheiden, Belgium).

## STAR METHODS

### Key Resource Table

STAR METHODS Key Resource Table
| REAGENT or RESOURCE | SOURCE | IDENTIFIER |
| --- | --- | --- |
| **<u>BIOLOGICAL SAMPLES</u>** |  |  |
| Human plasma samples | INNODIA consortium | N/A |
| **<u>CHEMICALS, PEPTIDES, AND RECOMBINANT PROTEINS</u>** |  |  |
| Ethanol, 200 proof | Sigma Aldrich | SHBL.2095/SHBK9944-5 |
| Library Dilution Buffer | HTG | 307900/308171 |
| **10 mM Tris pH 8.5 Elution Buffer** | QIAGEN | 163034340 |
| 2N NaOH | HTG | 308142 |
| 2N HCl | HTG | 307876 |
| Nuclease-Free Water (10x50 mL) | QIAGEN | 129114 |
| b-Mercaptoethanol | Sigma-Aldrich | M7522 |
| Tris EDTA buffer, for molecular biology, DNase, RNase, Proteasefree ready to use, pH 8.0 | Acros Organics | AC327345000 |
| **<u>CRITICAL COMMERCIAL ASSAYS</u>** |  |  |
| Plasma/Serum RNA Purification Mini Kit | NORGEN | 55000 |
| QIAseq miRNA Library Kit (96) | QIAGEN | 331505 |
| QIAseq miRNA 96 Index IL (96) | QIAGEN | 331565 |
| PhiX Control v3 | ILLUMINA | FC-110-3001 |
| NovaSeq 6000 SP Reagent Kit (100 cycles) | ILLUMINA | 20027464 |
| NovaSeq XP 2-Lane Kit | ILLUMINA | 20021664 |
| Thermo Qubit DSDNA HS Assay | LIFE<br>TECHNOLOGIES | Q32854 |
| Kit per DNA ad elevata intensità | AGILENT | 5067-4626 |
| NovaSeq 6000 S1 Reagent Kit v1.5 (100 cycles) | ILLUMINA | 20028319 |
| HTG Lysis Buffer | HTG | 307914 |
| MDx Proteinase K | Invitrogen | 2036707 |
| HTG Plasma Lysis Buffer | HTG | 308251 |
| EdgeSeq Drawer Reagents | HTG | 308312 |
| Human Brain RNA | Ambion | 2077661-A-2 |
| Hemo KlenTaq enzyme | New England<br>Biolabs | 10009977 |
| Deoxynucleotide (dNTP) Solution Set | New England<br>Biolabs | 10014675 |
| 5X Hemo KlenTaq Reaction Buffer | New England<br>Biolabs | 0031801 |
| PCR, H2O | HTG | 308124 |
| HTG Sequencing Tag Pack | HTG | 308065 |
| MIDx Ampure XP | HTG | 308183 |
| Bulk, Trizma Hydrochloride Solution (pH 8_0)<br>Dilution, HTG EdgeSeq | HTG | 308062 |
| MDX, I-tm (Md. BIO) | Lonza | RNBH 1710/RNBH3183 |
| KAPA Biosystems q PCR Kit | KAPA Biosystem | 005098 |
| MDx 12.5 PM PhiX | HTG | 308076 |
| HTI (Hybridization Buffer) | ILLUMINA | 20373892/20384289/<br>20391454 |
| Nextseq 500/550 Buffer cartridge v2 | ILLUMINA | 20399134/20383558 |
| NextSeq 500/550 High Output Reagent Cartridge v2 | ILLUMINA | 20396441 |
| NextSeq 500/550 High Output now cell Cartridge v2 | ILLUMINA | 20387266/20398769 |
| ILM Seq Read 1 Primer Mix Assy | HTG | 308246 |
| ILM Seq Index Primer Mix Assy | HTG | 308245 |
| TAQMAN PREAMP MASTER MIX (2X) | INVITROGEN | 4488593 |
| TAQMAN(R) MICRORNA RT KIT | INVITROGEN | 4366597 |
| TAQMAN MICRORNA ASSAY | INVITROGEN | cat.#4440887 ID 002332 |
| TAQMAN MICRORNA ASSAY | INVITROGEN | cat.#4440887 ID 000452 |
| TAQMAN MICRORNA ASSAY | INVITROGEN | cat.#4440887 ID 000572 |
| TAQMAN MICRORNA ASSAY | INVITROGEN | cat.#4440887 ID 000564 |
| ddPCR™ EvaGreen Supermix | BIORAD | 1864034 |
| ddPCR Smx for Probes (no dUTP), 5 x 1ml | BIORAD | 1863024 |
| DG8 Cartridges and Gaskets | BIORAD | 1864007 |
| ddPCR Plates 96-Well, Semi-Skirted (25 plates) | BIORAD | 12001925 |
| PIERCEABLE FOIL HEAT SEAL (no.100) | BIORAD | 1814040 |
| Droplet Gen Oil for Probes, 10 x 7 ml | BIORAD | 1863005 |
| **<u>SOFTWARE AND ALGORITHMS</u>** |  |  |
| Graphpad Prism v9.0.2 | GraphPad Software<br>(Boston, MA) | https://www.graphpad.com/ |
| 2100 Expert Software (vB.02.11) | Agilent | N/A; RRID:SCR_014466 |
| **Illumina Experiment Manager**<br>Software (v1.19.1) | ILLUMINA | N/A; RRID:SCR_021202 |
| BaseSpace v. 1.1.0.64 | ILLUMINA | N/A; RRID:SCR_011881 |
| GeneGlobe Data Analysis Center | QIAGEN | N/A |
| ExpressionSuite Software (v1.3) | THERMO FISHER | N/A; RRID:SCR_021095 |
| R/Bioconductor |  |  |
| **<u>OTHER</u>** |  |  |
| Vacutainer Safety-Lok butterflies | BD Biosciences | 367282 |
| NovaSeq Xp Flow Cell Dock | ILLUMINA | 20021663 |
| Vacutainer K2 EDTA | BD Biosciences | 368861 |
| DNA LoBind Tubes 0.5 mL | Eppendorf | 0030 108.035 |
| DNA LoBind Tubes 1.5 mL | Eppendorf | 0030 108.071 |
| DNA LoBind Tubes 2.0 mL | Eppendorf | 0030 108.058 |
| 96 Fast PCR Plate half skirt | SARSTEDT | 72.1981.202 |
| MicroAmp 96 Optical Adhesive Film 100 cover | Thermo Fisher | 4311971 |
| Strip 8 flat caps (Xtra-Clear) | STARLAB | I1400-0900 |
| StarTub Reagent Reservoir (PVC) | STARLAB | E2310-1000 |
| 96 PCR Plate without skirt | SARSTEDT | 72.1978.202 |
| SimpliAmp™ Thermal Cycler | LIFE<br>TECHNOLOGIES | A24811 |
| Qubit Assay Tube Set | Thermo Fisher | Q32856 |
| Centrifuge MiniSpin | Eppendorf | 5452000010 |
| Ministar centrifuge | VWR | 521-2319 |
| NovaSeq 6000 System | ILLUMINA | 20012850 |
| DynaMag-96 Side Magnet | INVITROGEN | 12331D |
| DynaMag-2 Magnet | INVITROGEN | 12321D |
| QUBIT 3.0 spectrofluorometer | INVITROGEN |  |
| 2100 Bioanalyzer Instrument | Agilent |  |
| **IKA MS3 S36 Agilent vortex IKA** | Agilent |  |
| HTG EdgeSeq Processor | HTG |  |
| HTG EdgeSeq Processor | HTG |  |
| SimpliAmp Thermal Cycler | LIFE<br>TECHNOLOGIES |  |
| StepOnePlus | APPLIED<br>BIOSYSTEM |  |
| NextSeq | ILLUMINA |  |

### Clinical Cohorts

#### INNODIA 100s cohort

For circulating small RNA sequencing analysis, an initial cohort composed of 115 individuals with newly diagnosed (<6 weeks, 4.5 ±1.5 weeks) type 1 diabetes were enrolled in INNODIA natural history study. T1DM individuals [all positive for at least one autoantibody (GADA, IA-2A, ZnT8A) and aged between 1-45 years] enrolled in this study were selected based on sample availability and even gender distribution (sex: 58F/57M; age at diagnosis: 12,4±7,7 years) (complete clinical characteristics in **Table 1 and TableS1**). T1DM individuals were followed-up to 24 months with programmed visits at 3- (V2), 6- (V3), 12- (V4) and 24-months (V5) after diagnosis. For circulating small RNAs study, we considered visits up to 12-months for the statistical association analysis with clinical parameters due to thei uncompleteness at V5 at the moment of small RNa measurement. Plasma samples for small RNAs sequencing were collected at baseline (V1, <6 weeks from diagnosis) through a standardised protocol^51^ adopted by all clinical sites involved in the multicentric consortium.

Specifically, blood samples were collected in K_2_ or K_3_-EDTA tubes and processed within 2 hours from blood draw. An initial centrifuge was performed at 1800g for 10 minutes at 15-25°C. Plasma was collected avoiding touching the white blood cells interphase and further centrifuged at 2000g for 20 minutes at 10°C to remove contaminant cells and platelets. Multiple 200 μL plasma aliquots were then stored at −80°C and then transferred to a central biobank located in Cambridge (UK) until final transfer to the analytical laboratory.

#### INNODIA 147s clinical cohort

An independent cohort composed by n=147 newly diagnosed (V1<6 weeks: 3.9±1.8 weeks) T1DM individuals [sex: 92/55 (M/F); age at diagnosis: 11,89 ± 7,86] was also enrolled in INNODIA consortium to perform the same analyses described for the 100s cohort. All individuals were positive for at least one autoantibody (GADA, IA-2A, ZnT8A) and aged between 1-45 years. All individuals were followed-up with programmed visits at 3- (V2), 6- (V3) and 12-months (V4) after diagnosis (**Table 2 ad Table S3**). Plasma samples at baseline visit 1 were collected following the SOP reported above.

### Ethics

The study followed the guidelines of the Declaration of Helsinki for research on human individuals, and the study was approved by the local ethical committees of the participating clinical sites (see **Table S2 and S4**). Participants gave written informed consent.

### HTG EdgeSeq miRNA whole transcriptome assay (Targeted-seq)

HTG EdgeSeq miRNA whole transcriptome assay method is an RNA extraction-free approach, and exploit quantitative nuclease protection assay (qNPA) chemistry with a subsequent Next Generation Sequencing (NGS) platform to allow semi-quantitative analysis of a panel of n=2102 targeted miRNAs (including n=13 housekeepings, n=5 negative process controls, n=1 positive process control and n=2083 targeted miRNAs) from 15 µl of plasma. In PCR-based library preparation, each sample is used as a template for PCR reactions for specially designed primers (tags), which share common sequences complementary to 5’-end and 3’-end “wing” sequences of the probes and common adapters required for cluster generation on NGS platform (Illumina NextSeq550). Libraries were prepared and cleaned-up with HTG EdgeSeq AMPure cleanup of Illumina Sequencing Libraries. Following libraries preparation, their concentration has been evaluated through HTG EdgeSeq KAPA Library quantification, and each library has been normalized and pooled using HTG EdgeSeq RUO library calculator. Then, pooled libraries were denatured in 2 N NaOH and sequenced (final concentration 4 pM) onto Illumina NextSeq550 platform (High Output kit v2 cat. FC-404-2005). Data were returned from the sequencer as demultiplexed FASTQ files. Resulting reads were aligned referring to miRbase v20 using HTG Parser software.

### QIAseq Small RNA sequencing (Untargeted-seq)

Total RNA extraction was performed from 200 µL of plasma through Serum/Plasma Norgen kit (cat. 55000, Thorold, ON L2V 4Y6, Canada). Small RNA-derived cDNA libraries were prepared using QiaSeq miRNA library kit (cat. 331505, Qiagen). QiaSeq strategy assign Unique Molecular Index (bound to reverse transcription primers) during reverse transcription step to every mature miRNA molecule, to enable unbiased and accurate miRNome-wide quantification of mature miRNAs by NGS. Then, libraries quality control (QC) was performed quantifying their concentration through QUBIT 3.0 spectrofluorometer (Qubit™ dsDNA HS Assay Kit, cat. Q32854, Thermofisher Scientific) and assessing their quality using capillary electrophoresis in Bioanalyzer 2100 (Agilent High Sensitivity DNA kit cat. 5067-4626, Thermofisher Scientific). High quality of libraries was evaluated considering electropherograms showing a peak comprised between 175 and 185 bp. Following QC, all libraries were normalized until 2 nM and pooled, denatured in 0.2 N NaOH and further sequenced (final concentration 175 pM) on Illumina NovaSeq 6000 platform [NovaSeq 6000 SP Reagent Kit (100 cycles) cat. 20027464, NovaSeq XP 2-Lane Kit cat. 20021664, Illumina] using the XP protocol applying 75x1 single reads.

Data were returned from BaseSpace Sequence Hub as demultiplexed FASTQ files. Resulting raw reads were deduplicated by leveraging Unique Molecular Identifiers (UMIs) present in the library, then mapped to miRbase v21 and piRNABank using QIAseq miRNA Quantification V1 Legacy pipeline from QIAGEN GeneGlobe Data Analysis Center portal (https://ngsdataanalysis2.qiagen.com/QIAseqmiRNA). Briefly, resulting reads were mapped referring to miRbase v21 and piRNABank using QIAGEN Gene Globe data analysis center software, which identified a wide repertoire of small RNA species e.g. piRNA (PIWI interacting RNA), tRFs (transfer RNA fragments), rRNA (ribosomal RNA), miRNA (microRNA).

All these procedures (samples collection and time, RNA extraction, Small RNAs library preparation and sequencing) were also conducted on 147s cohort as already described for 100s cohort, with minor modifications. In details, Small RNAs libraries were barcoded with unique dual indexes (UDI) (cat.# 331615 and cat.# 331625).

### Primary analysis of miRNA expression

Reads assigned to miRNAs were standardized into Counts Per Million (CPM) and filtered through *edgeR* package of R (*BioConductor*), maintaining only those miRNAs expressed in at least 70% of individuals with at least 15 CPM for HTG EdgeSeq and 10 CPM for QIAseq. Following low counts filtering, Median of Ratios normalization was performed through *DESeq2* package of R (*BioConductor*) and normalised counts were used for subsequent analyses. Consistently detected miRNAs in both sequencing platforms were selected, keeping only those having a positive Pearson correlation estimate between the two approaches (R>0 and p-value<0.05).

### Circulating miRNAs Unsupervised hierarchical clustering analysis

Unsupervised hierarchical clustering was independently applied to both miRNA expression datasets. The analyses were performed on log2 transformed data (after the addition of a pseudo-count), with *hclust* function of *stats* R package (complete-linkage agglomeration method and Pearson’s distance as distance metric). In order to determine the optimal cutting threshold silhouette method was applied.

Then, both dendrograms were split in two branches (*cutree* function of *stats* package of R with K=2) and named according to their size (Cluster A the major and Cluster B the minor). Only patients consistently identified as members of the same Cluster in both expression datasets were kept.

The association among patients’ clinical data and their branch membership was evaluated with logistic regression (glm function from *stats* package of R) and p-values < 0.05 were considered significant. The direct association between individual miRNA expression (log2-transformed, adding a pseudo-count) and each numerical clinical parameter was evaluated using Spearman’s correlation test. The same analysis was performed on Small RNAs sequencing dataset, obtained from the 147s cohort, by considering the same miRNAs used for the analyses in the 100s cohort.

### HLA genotyping

HLA typing was performed at v1 for n=107 out of n=109 T1DM individuals classified as Cluster A or Cluster B individuals through AXIOM Genotyping Array. HLA aplotype prevalence differences between Cluster A and Cluster B was assessed using chi-square test, considering significant P values ≤ 0.05.

### PBMC (cryopreserved) multi-dimensional flow cytometry (Multi-FACS) immunomics

Immunomics profile of peripheral blood from n=67 out of 115 individuals was investigated at baseline through Cytek Aurora flow cytofluorometer.

Samples were processed in five batches (between 12 to 16 samples per batch, consisting of a mixture of samples from each of the five INNODIA immune laboratories) together with two unrelated control samples in each batch using 0.54-2.8×10^6^ PBMCs per sample. PBMCs were first stained using Live/dead blue for 15 min at room temperature, washed with FACS buffer (PBS with 0.2% BSA and 2mM EDTA), and incubated with Fc receptor blocker (TruStain FcX Fc; BioLegend) for 10 min at room temperature. Without wash, samples were stained in a 37°C waterbath for 15 min using mastermix 1 (containing antibodies against CXCR3. CD117, CD294/CRTH2, and CD161). Samples were further stained in waterbath for 15 min using mastermix 2 (containing antibodies against CXCR5, ICOS, CCR7, and CCR6), followed by 30 min at room temperature using mastermix 3. Finally, samples were washed using FACS buffer, then fixed and resuspended in PBS containing 1% paraformaldehyde (Alfa Aesar). Single colour controls were made using PBMC for all colours except for CD294, CD117, CD161, and TCRgd where BD mouse or rat comp beads were used instead due to low cell expression. Single colour controls were subjected to the same buffer and fixed as the multi-colour stained samples. SpectroFlo QC beads were run daily and single colour controls were acquired in the reference library, which was subsequently used for live unmixing during sample acquisition on a Cytek Aurora cytometer. Flow data were analysed using FlowJo software using the Boolean gating scheme shown in **Figure S10**.

In order two find differences in the proportion of immune cell populations between the two cluster of patients (Cluster A, Cluster B), a Beta regression analysis was performed, using the *betareg* function (version 3.1.4)^94^. From the 109 patients assigned to the same Cluster by the two different sequencing platforms, 67 were also present in the immunomics cohort. Among these overlapping patients, n=48 belongs to Cluster A and n=19 to Cluster B. As first step of the analysis, proportions of immune cell population were transformed according to Smithson & Verkuilen^93^ to rescale the dependent variable in the interval (0,1), avoiding values of 0;1. Indeed, beta regression model cannot deal with values of 0 and 1. The Beta regression model was corrected for the time between blood draw and PBMCs isolation (same day vs. overnight). Immune cell populations with a *P* value≤0.05 related to the cluster were detected as significantly different in proportions between the two groups of patients. For representative purpose only, immune cell proportions were residualized for the time between blood draw and PBMCs isolation. For each immune cell population, a linear model was fit with the proportion as dependent variable and the time between blood draw and PBMCs isolation as independent variable. The residuals of the models represent the information on immune cell proportions that is not explianed from the timing of PBMC’s isolation.

### MiRNAs differential expression analysis

Normalised reads of the two sequencing datasets were used to detect any differentially expressed miRNAs between Cluster A and Cluster B groups in both sequencing platforms, accounting for age and sex as covariates with *DESeq2* package of R (*BioConductor*) using Wald test and Benjamini-Hoechberg adjusted *P* value ≤0.01 (considered as significant)

### Weighted miRNA correlation network analysis (WMCNA) for the identification of miRNA modules

Co-expression modules from the two different sequencing platforms were identified using the *WGCNA* package. Normalized expression values from the 248 common correlated miRNAs were transformed in log2 scale for the analysis (after the addition of one pseudo-count). Similarities between nodes were computed using the biweight midcorrelation, setting the max p-outliers parameter at 0.1 and using a weighted signed network. The following step was the identification of the Beta parameter (for both platforms) to compute adjacencies between nodes, by applying the approximate scale-free topology criterion. This criterion assumes that few highly connected nodes (hubs) link the rest of the less connected nodes to the system. Given the power-law distribution of the connectivity (sum of the adjacencies of a node with all the other nodes of the system), the goodness of the scale free-topology assumption for Beta values was measured through the R^2^ of the model regressing the log10 of probability of the connectivity and the log10 of the connectivity. High values of the R^2^ of the model are related to a straight line fitting the model, suggesting the assumption of the scale free topology. Moreover, the slope of the model should be close to −1. Candidate values of Beta ranging from 5 to 25 were manually inspected to choose the optimal ones for both platforms. Regarding the HTG-Seq platform, the scale free topology was met at a value of Beta equal to 24, which is the first value with an R^2^ close to 0.8 and with a slope of −0.82. On the other hand, regarding the Untargeted platform, the first value of Beta with an R² of the model of at least 0.8 was 11. However, the slope of the regression model for this value was very far from −1 (−0.40), suggesting that the number of nodes with a high connectivity does not decay as expected. Moreover, two values of Beta were very different for the two sequencing strategies (11 and 24) resulting in two networks with very different architectures. For this reason, the optimal value of Beta in the untargeted platform was determined as the first value of Beta with at least an R^2^ of the model of 0.8 and a regression slope of at least −0.7. The first value which satisfies these two criterions for the untargeted platform was Beta equal to 20. Once estimated the Beta parameters, similarities matrices were converted into adjacencies matrices by elevating similarities at the corresponding Beta value. The next step was the identification of the Topological Overlap Matrix (TOM) for both sequencing platforms from the adjacency matrices. The topological overlap of two nodes is a measure of similarity which defines how well the two nodes are interconnected. At this point the information from the two sequencing platforms, managed in a separate way during the previous phases of network construction, was merged in a consensus TOM. For the estimation of the consensus TOM, the two TOMs were first scaled at the 95^th^ quantile. The scaling step is crucial, because the consensus TOM was estimated as the minimum-wise component between the two TOMs. The consensus TOM, which contains the minimum-wise information about nodes interconnectivity from both platforms, was then transformed into a dissimilarity matrix (1-TOMcons). The *hclust* algorithm (hierarchical clustering), with average agglomeration method, was used to detect the modules, using the dissimilarity TOM as distance matrix. Minimum module size equal to 3 and deep split equal to 2 were set as parameters for the deepsplit function to cut the dendrogram for modules identification. At this point each miRNA was assigned to a module, with the grey one representing un-assigned miRNAs. The last step was the merging of very similar modules, to limit the redundancy in the information held. Modules EigenMiRNAs (MEs) were computed as the first principal component of the miRNAs expression values (normalized and log2 scaled) present in the module for both sequencing platforms. Similarity between MEs was computed with Pearson’s correlation coefficient, and then the dissimilarity was estimated and used for the hierarchical clustering of MEs. A cut height of 0.1 was used to merge closely related module. The final output of the *WGCNA* algorithm was a list of labels which identifies each miRNA as belonging to a specific module (after merging of closely related ones). Each module was summarized by a ME, computed as previously stated.

Once defined the modules through WMCNA, the aim of the analysis was the identification of a subset of miRNAs (n=5) for each module, which are the most representative. The centrality (hubness) of a miRNA within its module was defined using the intramodular connectivity as metric. The intramodular connectivity is the sum of the adjacencies of a miRNA with all the other miRNAs present in the module. The higher the intramodular connectivity, the higher the centrality of the node in the module. However, given the different values of Beta for the two sequencing platforms in the module’s estimation step (24 HTG-Seq, 20 Untargeted), the intramodular connectivity must be normalized in order to be comparable between them. Thus, for each module and sequencing platform, the intramodular connectivity of the node was divided by the maximum value of its own module. Then, the normalized intramodular connectivity of the nodes among sequencing platforms were summed up, and miRNAs were ranked based on this value. The 5 miRNAs with the highest values were defined as the most representatives for the module. Modules eigenMiRNAs of these subsets of most representative miRNAs were then computed for both sequencing platforms and correlated with clinical parameters using Spearman’s Rho correlation. Correlations with a p-value<0.05 were considered as significant.

### Droplet Digital PCR

Validation of selected miR-409-3p, miR-127-3p and miR-382-5p identified through differential expression analysis and WMCNA was performed through Custom Taqman reverse transcription and subsequent droplet digital PCR (ddPCR) detection.

In details, their expression was analysed in all plasma samples of 100s and 147s T1DM cohort using TaqMan miRNA assay primers (Life technologies, CA, USA) through a standardised protocol. RNA (the same used for small RNA sequencing) was reverse transcribed employing Custom RT primers pool and preamplified using Custom Preamp primers pool. Briefly, 5 µL each RT or TM primer was diluted in a total volume of 500 µL Tris-EDTA 1X and used for RT or preamplification reaction. Then, 3 μL of RNA were added to 6 μL of custom primers pool, 0.30 μL 100 mM dNTPs, 3 μL of 50 U/μL Multiscribe RT, 1.50 μL 10× RT Buffer, 0.19 μL 20 U/μL RNase Inhibitor and 1.01 μL H2O. The reaction product was incubated at 16 °C for 30 min, 42 °C for 30 min and then at 85 °C for 5 min. Afterwards, the synthesised cDNA was preamplified using Custom Preamp primer pool; the reaction included: 2.5 μL of cDNA from each sample, 12.5 μL 2× TaqMan Preamp Master Mix, 3.75 μL 10× Custom Preamp primers and 6.75 μL H_2_O. The reaction was incubated at 95 °C for 10 min, at 55 °C for 2 min and at 72 °C for 2 min, then for 12 cycles at 95 °C for 15 s and 60°C for 4 min and, finally, at 99 °C for 10 min. Then, droplet digital PCR was performed on a BioRad QX200 system using a Probes assay (BioRad, Mississauga, ON, Canada). Each PCR reaction contained 11 μL of QX200 super mix, 1.1 μL of each 20X TaqMan assay, 5.9 μL of H2O and 4 μL of template cDNA in a final volume of 22 μL. The PCR reactions were mixed, centrifuged briefly and 20 μL transferred into the sample well of a DG8™ cartridge. After adding 70 μL of QX200™ droplet generation oil into the oil wells, the cartridge was covered using a DG8™ gasket, and droplets generated using the QX200™ droplet generator. Droplets were carefully transferred into PCR plates using a multi-channel pipette and the plate sealed using PCR plate heat seal foil and the PX1™ PCR plate sealer. PCR was performed in a C1000 touch thermal cycler (BioRad, Mississauga, ON, Canada). The PCR protocol was 95°C for 10 min; 40 cycles of: 95°C for 30 s, optimal annealing temperature (56°C for miR-409-3p and miR-382-5p; 54°C for miR-127-3p; 98°C for 10 min; 4°C for 30 min. PCR plates were transferred into a QX200™ droplet reader to count positive and negative droplets. Thresholds to separate positive from negative droplets were set manually for each miRNA using the histogram function and reads analysed using QuantaSoft™ Analysis Pro software (Version 1.2, BioRad, Mississauga, ON, Canada).

### Quantification and statistical analysis

Sample size for circulating miRNAs analysis were determined according to our experience from previous works. Mann Whitney U test was performed between two groups when the variables did not follow a Gaussian distribution. Coefficient of Variation of miRNAs expression in Targeted- and Untargeted-seq was calculated on read counts using GraphPad 10.0.

Differences in clinical parameters between individuals belonging to Cluster A and Cluster B were determined with the univariate logistic regression using the glm function from the *stats* package in R software. Data were modelled using the Cluster A as dependent variable and the clinical parameter as independent variable. Clinical parameters with a P-value associated to the coefficient ≤0.05 were considered as significantly different between the clusters. The association between clinical parameters in each cluster of individuals was performed using a simple linear regression analysis, reporting β-values, R^2^ and *P*-values. Correlation analyses were performed using Spearman Rho Test or Pearson’s R test.

Statistical analyses were performed using R project (version 4.2.2) or GraphPad Prism (version 9 or 10).

