## Supplemental information for "A set of circulating microRNAs belonging to the 14q32 chromosomic locus identifies two clinically and phenotypically different subgroups of individuals with recent onset Stage 3 type 1 diabetes"

*Title page*

1- Diabetes Unit, Department of Medicine, Surgery and Neurosciences, University of Siena, Siena, Italy
2- Fondazione Umberto Di Mario ONLUS c/o Toscana Life Science, Siena, Italy.
3- Tuscany Centre for Precision Medicine (CReMeP), Siena, Italy.
4- Diabetes Research Institute, Leonard Miller School of Medicine, University of Miami, FL, USA
5- Department of Diabetes Immunology, Arthur Riggs Diabetes and Metabolism Research Institute, Beckman Research Institute, City of Hope, Duarte, CA, USA.
6- Center for Diabetes and Metabolic Diseases and the Wells Center for Pediatric Research, Indiana University School of Medicine, Indianapolis, IN, USA.
7- Katholieke Universiteit Leuven/Universitaire Ziekenhuizen, Leuven, Belgium.
8- Department of Immunobiology, King's College London, School of Immunology and Microbial Sciences, London, UK.
9- Immunology & Inflammation Research Therapeutic Area, Sanofi, Boston, MA, USA.
10- Lead Contact

*Shared First co-authorship

**^§^** INNODIA: “Innovative approaches to understanding and arresting type 1 diabetes”- List

of contributors included with submission.

Contact info:

Prof. Francesco Dotta

Diabetes Unit, Dept. of Medicine, Surgery and Neurosciences,

University of Siena, Siena, Italy

**SUPPLEMENTAL FIGURES**

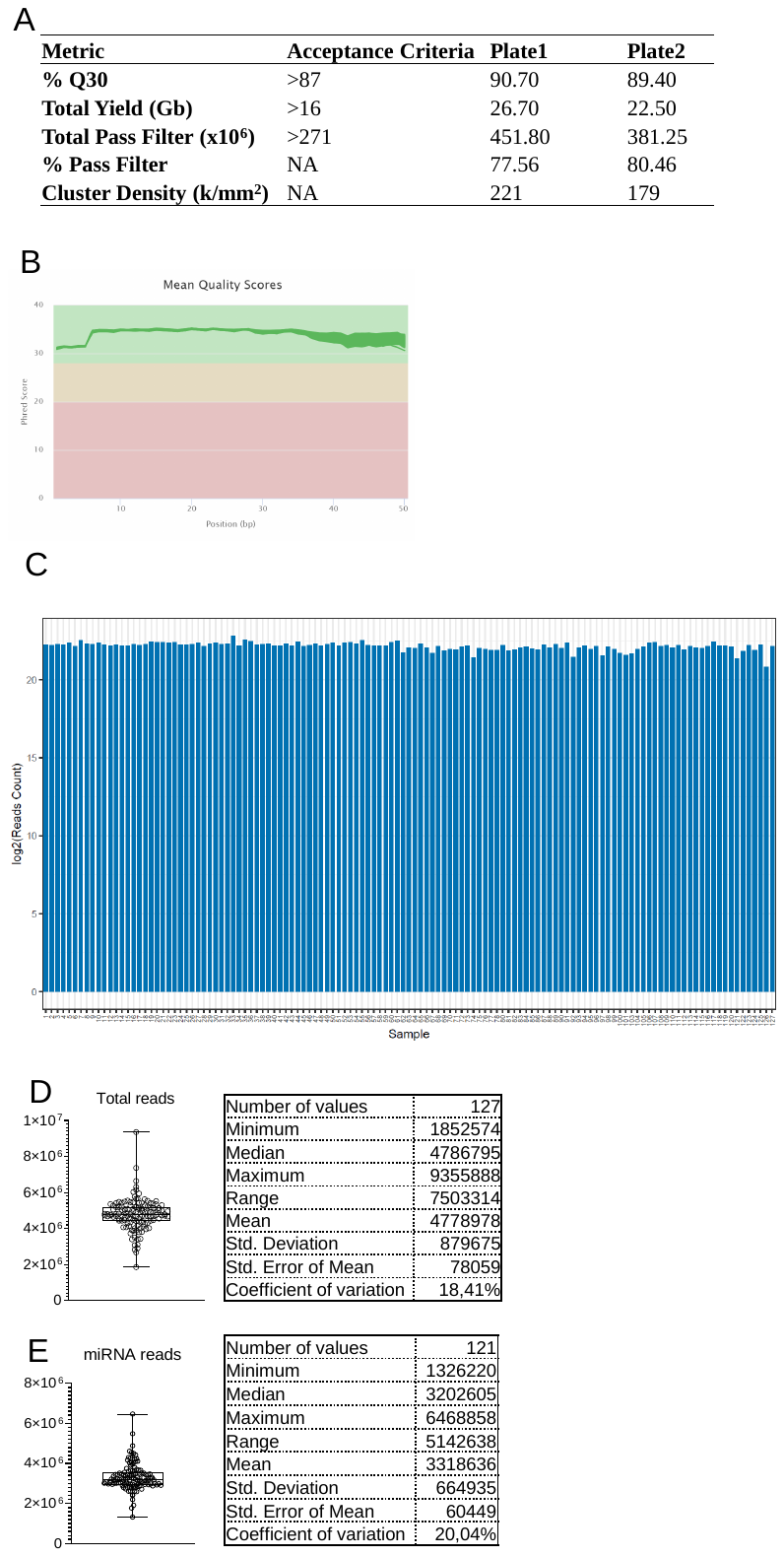

**Figure S1. Sequencing quality metrics of HTG miRNA Edge seq or Targeted-seq.** (A) Main sequencing metrics of the sequencing run executed on n=127 samples using Illumina NextSeq550 sequencer. Sequencing metrics related to reads quality score (%QC30), total yield reported as gigabases (Gb), total reads pass filter, percentage pass filter and cluster density are reported in comparison to Illumina standards quality cut-offs. (B) Phred Score graph reporting the mean quality for each nucleotide of the reads sequenced. Sequencing cycles are reported on x-axis while per base average Phred score is reported on y-axis. (C) Bar plot reporting total reads count obtained for each sample sequenced through HTG-Edge seq platform. (C) Box-and-whisker plot (min-to max) reporting Total Reads per sample obtained through HTG-Edge seq platform. Total reads and (**D**) miRNA reads (**E**) per samples obtained through small RNA-seq; descriptive statistics are reported on the right for each analysis. Each dot represents a sample; mean along with min-to-max error bars are reported both in (**D**) and (**E**).

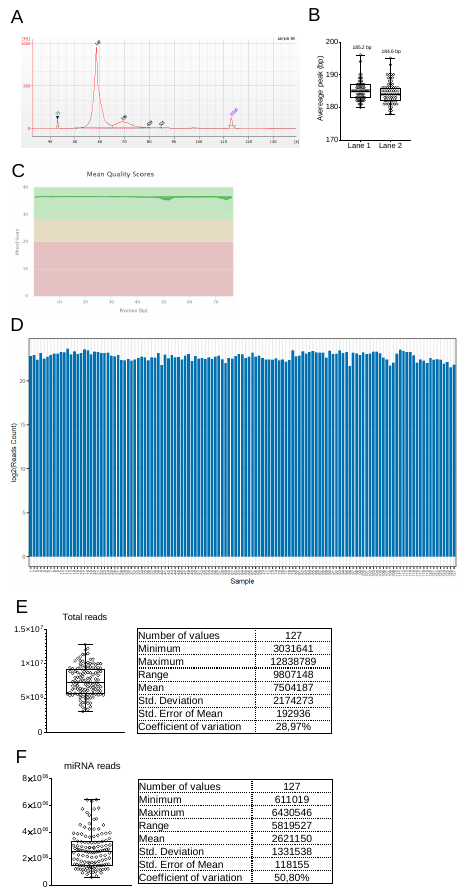

**Figure S2. Quality check of QiaSeq miRNA Libraries and Small RNA-seq or untargeted-seq sequencing metrics**. (**A**) Representative capillary electrophoresis quality control analysis on a cDNA library generated from plasma small RNAs through Qiaseq miRNA Library preparation kit. The capillary electrophoresis was performed using Agilent 2100 Bioanalyzer. The electropherogram shows a major peak at 180 bp, as expected. (**B**) Box plot reporting the average peak of each sequenced cDNA library analysed using capillary electrophoresis; each dot represents a library. Values are shown as base pair (bp) length of the average peak. Results are reported separately for each sequencing lane used on NovaSeq 6000. (**C**) Phred Score graph reporting the mean quality for each nucleotide of the reads sequenced. Sequencing cycles are reported on x-axis while per base average Phred score is reported on y-axis. (**D**) Bar plot reporting total reads counts of all samples analysed (n=127), obtained through small RNA-seq or untargeted-seq. Box-and-whisker plot (min-to max) reporting (**E**) total reads and (**F**) miRNA reads per samples obtained through small RNA-seq; descriptive statistics are reported on the right for each analysis. Each dot represents a sample; mean along with min-to-max error bars are reported both in (**D**) and (**E**).

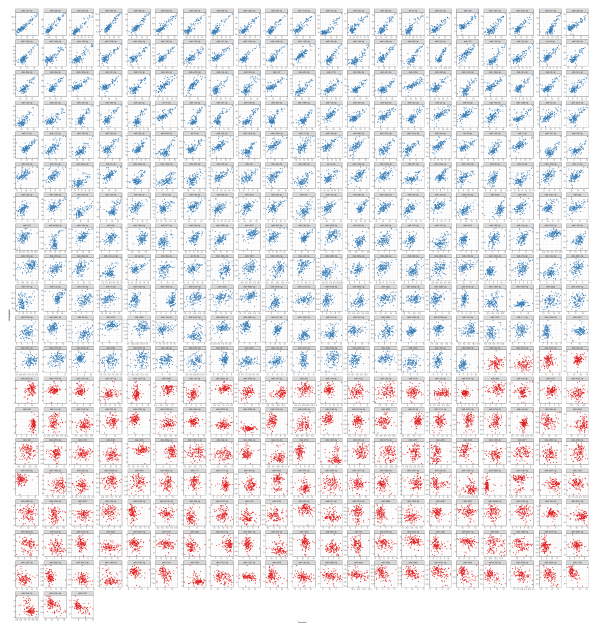

**Figure S3.** Concordant miRNAs selection based on Pearson R Estimates (R>0, *P*≤0.05) between targeted and untargeted-seq datasets. Each graph reports a scatter plot of the expression levels of the common set of detected miRNAs (n=402) in both platforms (x = targeted-seq; y = untargeted-seq). Scatter plots are arranged from the most to the least concordant miRNA between the two datasets. Blue = common-concordant miRNAs (n=248); red = common-discordant miRNAs (n=154). Each dot represents a T1DM subject (n=115).

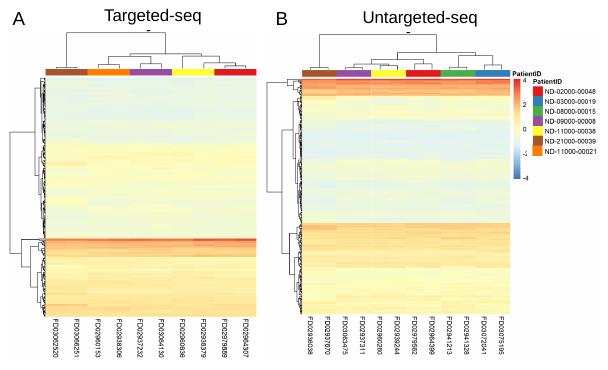

**Figure S4. Unsupervised hierarchical clustering of technical replicates.** Unsupervised hierarchical clustering analysis was performed on technical replicate sample pairs in both targeted-seq (**A**) and untargeted-seq (**B**) platforms. The hierarchical clustering shows different color-coded technical replicates. In both platforms, the technical replicate sample pairs clustered together. The hierarchical clustering analysis was performed on the common and concordant miRNA set (n=248; y axis) and different plasma aliquots from the same patients (x axis). The scale color ranges from red to blue, representing the scaled expression values for each detected miRNA.

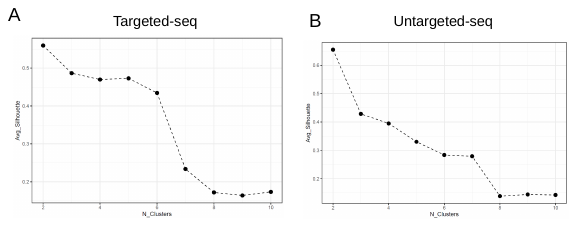

**Figure S5. Determination of the optimal number of T1DM clusters in Targeted and Untargeted-seq.** The silhouette method measures the similarities of each T1DM subject to its own cluster respect to the other clusters, based on a distance value (calculated as 1-Pearson R). For each sequencing platform the average silhouette value of T1DM subjects for *k* clusters [2 to 10] was calculated. The value of *k* is reported on *x*-axis while average silhouette values are reported on *y*-axis. It was chosen *k* = 2, as it maximizes the silhouette value both in targeted- and untargeted-seq.

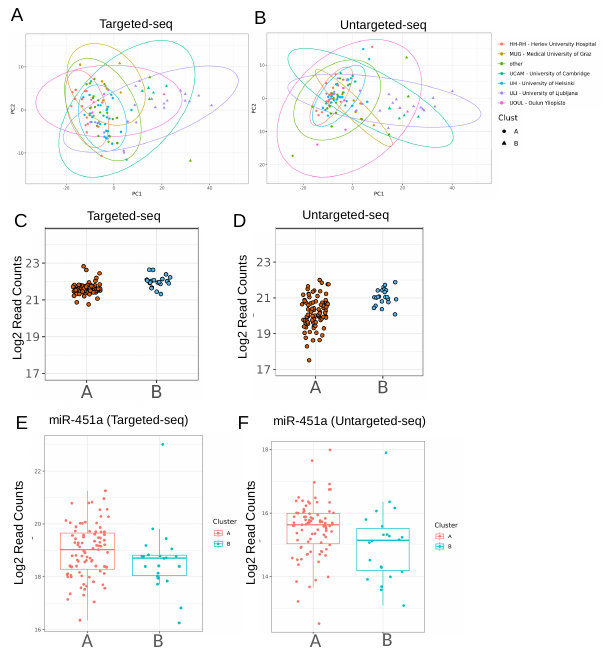

**Figure S6. Analysis of putative confounding variables in the determination of clustering of T1DM subjects.** (A-B) Principal Component Analysis (PCA) of miRNA expression in targeted-seq (A) and untargeted-seq (B), displaying the clustering of samples and indicating their membership in either Cluster-A or Cluster-B, as well as their clinical site of origin. Elliptical groupings of samples from each clinical site are depicted and color-coded for easy reference. (C-D) Total miRNA read counts in Cluster-A and Cluster-B for targeted-seq (C) and untargeted-seq (D). Each dot represents the total miRNA read count per sample. The data are expressed as log2 of total miRNA counts. (E-F) Bar plots presenting the expression of the hemolysis indicator miRNA miR-451a in Cluster-A and Cluster-B. No significant difference was observed between Cluster-A and Cluster-B in targeted-seq (E) and untargeted-seq (F). The data are depicted as individual dots for each sample, along with the median, first and third quartile. Statistical analysis was performed using the non-parametric Mann-Whitney test (P ≤ 0.05).

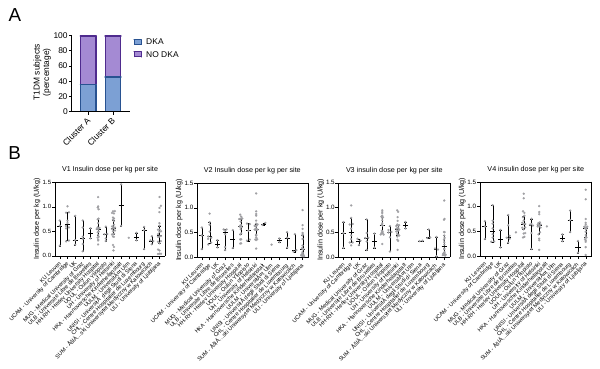

**Figure S7**. (**A**) Diabetic ketoacidosis rate at T1DM onset between Cluster-A and Cluster-B. Data are reported as a percentage of the total T1DM subjects included in each respective cluster. Statistical analysis was performed using a chi-square test (P ≤ 0.05). (**B**) Insulin daily dose per kg at baseline (V1) and follow-up visits (V2, V3, V4) for each T1DM subject included in the 100s cohort and subdivided based on the clinical site of recruitment. Available insulin dose values are reported as insulin units per day per kg for each T1DM subject. For representation purposes, three different satellite clinical centers in the UK were reported together as UK.

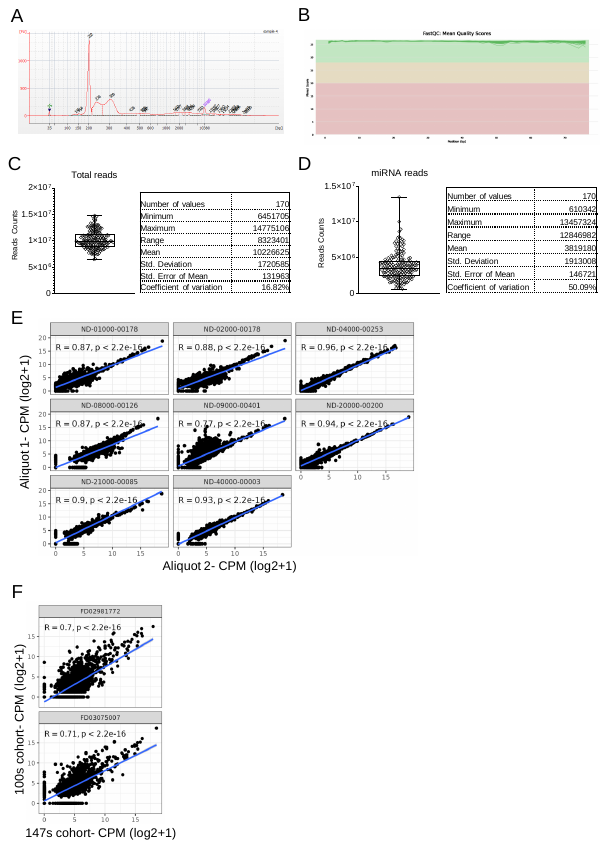

**Figure S8. Quality Check of Qiaseq miRNA Libraries and Small RNA-seq or untargeted-seq sequencing metrics**. (**A**) Representative capillary electrophoresis quality control analysis of a cDNA library generated from plasma small RNAs using the Qiaseq miRNA Library preparation kit. The analysis was performed using an Agilent 2100 Bioanalyzer. The electropherogram demonstrates a major peak at 200 bp, as expected. (**B**) Phred Score graph displaying the Q30 scores for each nucleotide of the sequenced reads. The x-axis represents the position in base pairs (bp), while the y-axis represents the total Phred score. (**C**) Box plot showing the total reads obtained through small RNA-seq. (**D**) Box plot presenting the miRNA reads per sample obtained through small RNA-seq. Descriptive statistics are provided on the right side for each analysis. (**E**) Correlation analysis between internal replicate sample pairs (n=8) representing two different aliquots (aliquot-1, aliquot-2) from the same subject's plasma sample. The subject ID, along with the corresponding Pearson R and P values, are indicated for each correlation. Each dot represents an individual miRNA. Expression values are reported as log2 Counts Per Million (CPM). The statistical analysis was performed using the Pearson R correlation test. (**F**) Correlation analysis of inter-cohort samples (100s vs 147s cohort). A total of n=2 plasma sample aliquots were run in both the 100s and 147s cohorts. The correlation was performed on commonly detected miRNAs (n=226) for each inter-cohort sample pair using the Pearson R correlation test (p ≤ 0.05). Each dot represents an individual miRNA commonly detected in both the 100s and 147s cohorts. Expression values are reported as log2 Counts Per Million (CPM).

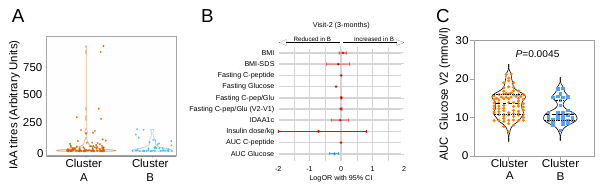

**Figure S9. Clinical characteristics associated with Cluster-A and Cluster-B T1DM subjects in the 147s cohort.** (**A**) IAA titers measured at baseline (V1) in T1DM subjects of the 147s cohort subdivided into Cluster-A and Cluster-B. (**B**) Forest plot presenting the effect estimates (represented by single dots) and 95% confidence intervals (indicated by bars) for Cluster-B across selected clinical variables collected at V2. The effects of Cluster-B are presented as log odds ratio using univariate logistic regression analysis. Blue bars indicate statistically significant effects (P ≤ 0.05). (**C**) Area under the curve of glucose (calculated using the trapezoidal rule) measured during the mixed meal tolerance test (MMTT) at visit 2 in T1DM subjects of the 147s cohort, subdivided into Cluster-A and Cluster-B. Statistical analysis was performed using the Mann-Whitney test (P ≤ 0.05).

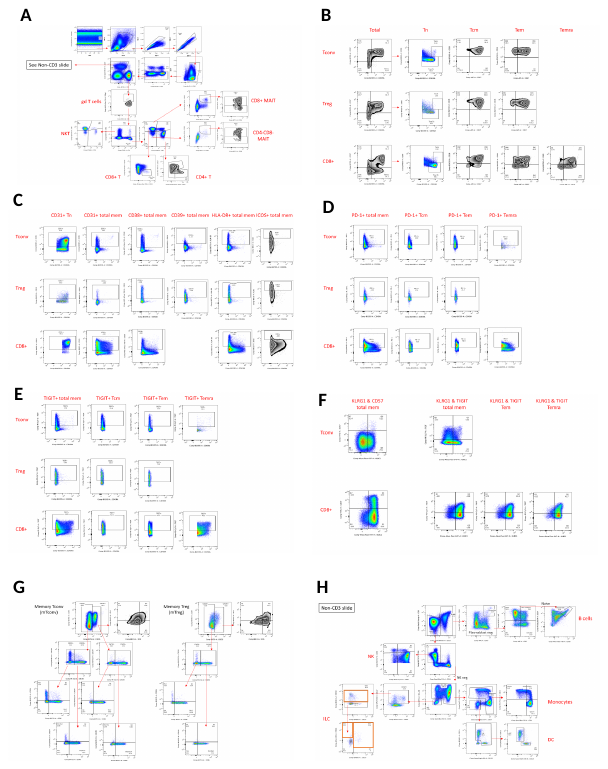

**Figure S10. High-Parameter Flow cytometry Boolean gating strategy to identify the main circulating immune cell subsets in peripheral blood of T1DM individuals at baseline visit.**

**SUPPLEMENTAL TABLES**

**Table S1A. Baseline (visit 1) and follow-up (visit 2, visit 3 and visit 4) clinical characteristics of 100s cohort T1DM subjects population.** Mean values ± standard deviation are reported for continuous variables; n or n% for categorical values. Number of T1DM subjects with available measurements for each specific variable is reported in brackets. BMI-SDS is exclusively reported for T1DM subjects below 18 years of age (n=16) at diagnosis.

| **100s cohort T1DM subjects demographics** | **Visit 1**  **(baseline, 0 months)** | **Visit 2**  **(3 months)** | **Visit 3**  **(6 months)** | **Visit 4**  **(12 months)** |
| --- | --- | --- | --- | --- |
| Age (years) | 12.5 ± 7.7 [115] | 12.5 ± 7.5 [114] | 12.97 ± 7.70 [115] | 13.46 ± 7.53 [100] |
| Sex (Female/Male) | 58/57 |  |  |  |
| BMI (Kg/m^2^) | 23.1 ± 2.8 [16] | 24.0 ± 2.66 [16] | 24.5 ± 2.9 [18] | 24.4 ± 2.9 [17] |
| BMI_SDS | 0.15 ± 1.09 [99] | 0.18 ± 1.06 [96] | 0.01 ± 1.10 [94] | 0.13 ± 1.03 [83] |
| Disease duration (weeks) | 4.1 ± 1.5 [115] | 14.3 ± 5.16 [114] | 27.0 ± 2.29 [115] | 53.0 ± 2.5 [100] |
| Insulin dose (Units/day/kg) | 0.52 ± 0.27 [113] | 0.46 ± 0.25 [106] | 0.47 ± 0.25 [108] | 0.57 ± 0.27 [93] |
| Fasting C-peptide (pmol/L) | 277.6 ± 203.5 [115] | 316.1 ± 224.6 [103] | 290.3 ± 214.5 [103] | 235.6 ± 215.8 [90] |
| HbA1c (mmol/mol) | 77.4 ± 19.4 [112] | 48.13 ± 9.75 [108] | 50.8 ± 11.4 [107] | 53.9 ± 12.4 [97] |
| DKA (No/Yes) | 73 / 41 [114] | ─ | ─ | ─ |
| GAD65 (% positive) | 76 [115] | n/a | n/a | 71,1 [97] |
| IA2A (% positive) | 72,1 [115] | n/a | n/a | 70,1 [97] |
| IAA (% positive) | 76 [115] | n/a | n/a | 100 [97] |
| ZnT8A (% positive) | 66 [115] | n/a | n/a | 54,7 [97] |

**Table S1B**. **Baseline (visit 1) clinical characteristics of 100s cohort T1DM subjects population in the two identified clusters (Cluster A and Cluster B).** Mean values ± standard deviation are reported for continuous variables; n or n% for categorical values. Number of T1DM subjects with available measurements for each specific variable is reported in brackets. BMI-SDS is exclusively reported for T1DM subjects below 18 years of age at diagnosis.

| **100s cohort T1DM subjects demographics** | **Cluster A [n=87]** | **Cluster B [n=22]** |
| --- | --- | --- |
| Age (years) | 12.49 ± 7.64 [87] | 13.0 ± 8.86 [22] |
| Sex (Female/Male) | 42/45 [87] | 14/8 [22] |
| BMI (Kg/m^2^) | 18.83 ± 3.83 [87] | 18.22 ± 3.94 [22] |
| BMI_SDS | 0.3 ± 1.09 [87] | 0.09 ± 1.11 [22] |
| Disease duration (weeks) | 4.27 ± 1.41 [87] | 3.32 ± 1.83 [22] |
| Insulin dose (Units/day/kg) | 0.54 ± 0.27 [85] | 0.48 ± 0.28 [22] |
| Fasting C-peptide (pmol/L) | 290.89 ± 213.6 [87] | 245.51 ± 160.69 [22] |
| HbA1c (mmol/mol) | 75.11 ± 17.19 [85] | 89.36 ± 21.54 [21] |
| DKA (No/Yes) | 56/30 [86] | 12/10 [22] |
| GAD65 (% positive) | 77,01 [67] | 77,27 [17] |
| IA2A (% positive) | 74,71 [65] | 68,18 [15] |
| IAA (% positive) | 79,31 [69] | 63,64 [14] |
| ZnT8A (% positive) | 66,67 [58] | 63,64 [14] |
| Age Stratification (<7/≥7 yo) | 14/73 [87] | 3/19 [22] |
| Age Stratification (<7/≥7 yo) ratio | 1:5.21 | 1:6.3 |

**Table S2. Number of T1DM subjects from 100s cohort per clinical site (n=16) in each visit.**

| **Clinical Site** | **Visit 1**  **(baseline)** | **Visit 2**  **(3 months)** | **Visit 3**  **(6 months)** | **Visit 4**  **(12 months)** |
| --- | --- | --- | --- | --- |
| CHL - Centre Hospitalier de Luxembourg - Luxembourg | 3 | 3 | 3 | 3 |
| HH-RH - Herlev University Hospital - Copenhagen, Denmark | 20 | 20 | 20 | 19 |
| HKA - Hannoversche Kinderheilanstalt - Hannover, Germany | 2 | 2 | 2 | 2 |
| KU Leuven - Leuven, Belgium | 3 | 3 | 3 | 3 |
| MUG - Medical University of Graz - Graz, Austria | 6 | 6 | 6 | 6 |
| SUM - Śląski Uniwersytet Medyczny w Katowicach- Katovice, Poland | 4 | 4 | 4 | 2 |
| UCAM - University of Cambridge- Cambridge, UK | 10 | 10 | 10 | 9 |
| UH - University of Helsinki- Helsinki, Finland | 28 | 28 | 28 | 20 |
| UK - Barts Health NHS Trust- London, UK | 3 | 3 | 3 | 3 |
| UK - Norfolk and Norwich- Norwich Norfolk, UK | 1 | 1 | 1 | 1 |
| UK - North West Anglia- Peterborough, UK | 1 | 1 | 1 | 0 |
| ULB - Universite Libre de Bruxelles- Bruxelles, Belgium | 3 | 3 | 3 | 2 |
| ULI - University of Ljubljana- Ljubljana, Slovenia | 22 | 22 | 22 | 21 |
| UNISI - Università degli Studi di Siena- Siena, Italy | 2 | 2 | 2 | 2 |
| UOUL - Oulun Yliopisto- Oulu, Finland | 6 | 5 | 6 | 6 |
| UULM - Universität Ulm- Ulm, Germany | 1 | 1 | 1 | 1 |
| TOTAL | 115 | 114 | 115 | 100 |

**Table S3A Baseline (visit 1) and follow-up (visit 2, visit 3 and visit 4) clinical characteristics of 147s cohort T1DM subjects population.** Mean values ± standard deviation are reported for continuous variables; n or n% for categorical values. Number of T1DM subjects with available measurements/information for each specific variable is reported in brackets. BMI-SDS is exclusively reported for T1DM subjects below 18 years of age at the moment of diagnosis.

| **147s cohort subject demographics** | **Visit 1** | **Visit 2** | **Visit 3** | **Visit 4** |
| --- | --- | --- | --- | --- |
| Age(years) | 12,03 ± 7,82 [147] | 12,21 ± 7,83 [146] | 12,48 ± 7,86 [145] | 12,98 ± 7,79 [147] |
| Sex (Female/Male) | 55/92 [147] | 55/91 [146] | 55/90 [145] | 55/92 [147] |
| BMI (Kg/m2) | 22,26 ± 2,99 [21] | 22,52 ± 3,31 [18] | 22,72 ± 3,53 [21] | 23,29 ± 3,48 [26] |
| BMI_SDS | 0,38 ± 1,12 [126] | 0,38 ± 1,08 [115] | 0,34 ± 1,07 [121] | 0,45 ± 1,1 [120] |
| Disease duration (weeks) | 3,91 ± 1,76 [142] | 14,83 ± 4,48 [141] | 27,91 ± 5,01 [140] | 53,76 ± 4,84 [142] |
| Insulin dose (units/day/kg) | 0,59 ± 0,40 [144] | 0,45 ± 0,26 [130] | 0,48 ± 0,24 [139] | 0,57 ± 0,25 [143] |
| Fasting C-peptide (pmol/L) | 270,03 ± 194,79 [145] | 319,90 ± 227,61 [129] | 278,78 ± 202,28 [131] | 224,19 ± 214,55 [121] |
| DKA (No/Yes) | 83/57 [140] | / | / | / |
| HbA1c (mmol/mol) | 76,01 ± 19,26 [143] | 48,39 ± 8,83 [133] | 50,22 ± 9,62 [139] | 53,33 ± 10,64 [143] |
| IAA (% positive) | 77,55 [114] | n/a | n/a | 97,96 [144] |
| IA2A (% positive) | 78,23 [115] | n/a | n/a | 73,47 [108] |
| GAD65 (% positive) | 77,55 [114] | n/a | n/a | 72,79 [107] |
| ZnT8A (% positive) | 70,75[104] | n/a | n/a | 66,67 [98] |

**Table S3B. Baseline (visit 1) clinical characteristics of 147s cohort T1DM subjects population in the two identified clusters (Cluster A and Cluster B).** Mean values ± standard deviation are reported for continuous variables; number or percentage for categorical values. Number of T1DM subjects with available measurements for each specific variable is reported in brackets. BMI-SDS is exclusively reported for T1DM subjects below 18 years of age (n=16) at the moment of diagnosis

| **147s cohort T1DM subjects demographics** | **Cluster A [n=105]** | **Cluster B [n=42]** |
| --- | --- | --- |
| Age (years) | 11.32 ± 7.29 [105] | 13.79 ± 8.86 [42] |
| Sex (Female/Male) | 45/60 [105] | 10/32 [42] |
| BMI (Kg/m^2^) | 18.65 ± 3.25 [105] | 19.47 ± 3.51 [42] |
| BMI_SDS | 0.39 ± 1.16 [105] | 0.4 ± 0.89 [42] |
| Disease duration (weeks) | 3.8 ± 1.75 [102] | 4.21 ± 1.76 [40] |
| Insulin dose (Units/day/kg) | 0.62 ± 0.44 [103] | 0.53 ± 0.29 [41] |
| Fasting C-peptide (pmol/L) | 272.52 ± 196.88 [105] | 263.5 ± 191.51 [40] |
| HbA1c (mmol/mol) | 76.06 ± 19.19 [103] | 75.87 ± 19.67 [40] |
| DKA (No/Yes) | 56/45 [101] | 28/12 [40] |
| GAD65 (% positive) | 76,19 [80] | 80,95 [34] |
| IA2A (% positive) | 76,19 [80] | 83,33 [35] |
| IAA (% positive) | 78,1 [82] | 76,19 [32] |
| ZnT8A (% positive) | 74,29 [78] | 61,9 [26] |
| Age stratification (<7/≥7 yo) | 23/82 [105] | 7/35 [42] |
| Age stratification (<7/≥7 yo) ratio | 1:3.5 | 1:5.0 |

**Table S4. Number of T1DM subjects from 147s cohort per clinical site (n=15) in each visit.**

| **Clinical Site** | **Visit 1** | **Visit 2** | **Visit 3** | **Visit 4** |
| --- | --- | --- | --- | --- |
| CHL - Centre Hospitalier de Luxembourg, Luxembourg | 19 | 19 | 19 | 19 |
| HH-RH - Herlev University Hospital, Copenhagen-Denmark | 8 | 8 | 8 | 8 |
| HKA - Hannoversche Kinderheilanstalt, Hannover-Germany | 2 | 2 | 2 | 2 |
| IT- Ospedale Pediatrico Bambino Gesù, Roma- Italy | 5 | 5 | 5 | 5 |
| KU Leuven, Leuven- Belgium | 9 | 9 | 9 | 9 |
| MUG - Medical University of Graz, Graz- Austria | 11 | 11 | 11 | 11 |
| SUM - Slaski Uniwersytet Medyczny w Katowicach, Katovice, Poland | 9 | 9 | 8 | 9 |
| UCAM - University of Cambridge, Cambridge- UK | 6 | 5 | 5 | 6 |
| UH - University of Helsinki, Helsinki- Finland | 39 | 39 | 39 | 39 |
| UK- Barts Health NHS Trust, London- UK | 2 | 2 | 2 | 2 |
| UK - Leicester Royal Infirmary, Leicester- UK | 2 | 2 | 2 | 2 |
| ULB - Universitè Libre de Bruxelles, Bruxelles- Belgium | 6 | 6 | 6 | 6 |
| ULI - University of Ljubljana, Ljiubljana- Slovenia | 16 | 16 | 16 | 16 |
| UNISI - Universita degli Studi di Siena, Siena-Italy | 4 | 4 | 4 | 4 |
| UOUL - Oulun Yliopisto, Oulu, Finland | 9 | 9 | 9 | 9 |

**Table S5. Numerousness of available clinical parameters for each visit in Cluster-A and Cluster-B T1D subjects of 100s and 147s cohort**

|  | **Cohort** | | | | | |
| --- | --- | --- | --- | --- | --- | --- |
|  | **100s** | |  |  | **147s** | |
| **Clinical Parameter** | **Cluster-A (n=87)** | **Cluster-B (n=22)** |  |  | **Cluster-A (n=105)** | **Cluster-B (n=42)** |
| v1_BMI | 87 | 22 |  |  | 105 | 42 |
| v1_BMI_SDS | 87 | 22 |  |  | 105 | 42 |
| v1_IDAA1c | 83 | 21 |  |  | 101 | 39 |
| v1_age_at_visit | 87 | 22 |  |  | 105 | 42 |
| v1_fasting_c_pep_glu_ratio | 87 | 22 |  |  | 104 | 39 |
| v1_fasting_c_pep_result | 87 | 22 |  |  | 105 | 40 |
| v1_hbalc_result | 85 | 21 |  |  | 103 | 40 |
| v1_insulin_dose_per_kg | 85 | 22 |  |  | 103 | 41 |
| v1_v2_fasting_c_pep_glu_ratio_change | 72 | 18 |  |  | 83 | 29 |
| v1_weeks_from_diagnosis | 87 | 22 |  |  | 102 | 40 |
| v2_BMI | 84 | 22 |  |  | 97 | 36 |
| v2_BMI_SDS | 84 | 22 |  |  | 97 | 36 |
| v2_IDAA1c | 77 | 20 |  |  | 91 | 35 |
| v2_age_at_visit | 86 | 22 |  |  | 105 | 41 |
| v2_fasting_c_pep | 78 | 19 |  |  | 94 | 35 |
| v2_fasting_c_pep_glu_ratio | 72 | 18 |  |  | 84 | 31 |
| v2_fasting_c_pep_result | 78 | 19 |  |  | 94 | 35 |
| v2_fasting_glucose | 74 | 20 |  |  | 87 | 33 |
| v2_hbalc_result | 81 | 21 |  |  | 96 | 37 |
| v2_insulin_dose_per_kg | 80 | 20 |  |  | 95 | 35 |
| v2_mmtt_auc_c_pep | 73 | 20 |  |  | 81 | 30 |
| v2_mmtt_auc_glucose | 72 | 20 |  |  | 80 | 31 |
| v2_v3_fasting_c_pep_glu_ratio_change | 65 | 16 |  |  | 76 | 29 |
| v2_v3_mmtt_auc_change | 63 | 18 |  |  | 73 | 28 |
| v2_weeks_from_diagnosis | 86 | 22 |  |  | 102 | 39 |
| v3_BMI | 85 | 22 |  |  | 102 | 40 |
| v3_BMI_SDS | 84 | 22 |  |  | 102 | 40 |
| v3_IDAA1c | 76 | 20 |  |  | 98 | 38 |
| v3_age_at_visit | 87 | 22 |  |  | 104 | 41 |
| v3_fasting_c_pep | 76 | 21 |  |  | 96 | 35 |
| v3_fasting_c_pep_glu_ratio | 69 | 19 |  |  | 86 | 31 |
| v3_fasting_c_pep_result | 76 | 21 |  |  | 96 | 35 |
| v3_fasting_glucose | 73 | 20 |  |  | 90 | 33 |
| v3_hbalc_result | 79 | 22 |  |  | 99 | 40 |
| v3_insulin_dose_per_kg | 82 | 20 |  |  | 101 | 38 |
| v3_mmtt_auc_c_pep | 68 | 20 |  |  | 86 | 31 |
| v3_mmtt_auc_glucose | 67 | 19 |  |  | 86 | 32 |
| v3_v4_fasting_c_pep_glu_ratio_change | 57 | 16 |  |  | 72 | 26 |
| v3_v4_mmtt_auc_change | 53 | 16 |  |  | 70 | 24 |
| v3_weeks_from_diagnosis | 87 | 22 |  |  | 101 | 39 |
| v4_BMI | 75 | 19 |  |  | 105 | 42 |
| v4_BMI_SDS | 74 | 19 |  |  | 103 | 42 |
| v4_IDAA1c | 67 | 18 |  |  | 100 | 39 |
| v4_age_at_visit | 75 | 19 |  |  | 105 | 42 |
| v4_fasting_c_pep | 66 | 19 |  |  | 86 | 35 |
| v4_fasting_c_pep_glu_ratio | 63 | 19 |  |  | 78 | 30 |
| v4_fasting_c_pep_result | 66 | 19 |  |  | 86 | 35 |
| v4_fasting_glucose | 69 | 19 |  |  | 92 | 36 |
| v4_hba1c_result | 73 | 18 |  |  | 101 | 42 |
| v4_insulin_dose_per_kg | 69 | 19 |  |  | 104 | 39 |
| v4_mmtt_auc_c_pep | 60 | 19 |  |  | 76 | 30 |
| v4_mmtt_auc_glucose | 62 | 19 |  |  | 85 | 30 |
| v4_weeks_from_diagnosis | 75 | 19 |  |  | 102 | 40 |
